# Accelerated metabolomic ageing (MileAge) in mid-life predicts incident vascular, unspecified and all-cause dementia

**DOI:** 10.1101/2025.05.09.25327305

**Authors:** Julian Mutz, Lachlan Gilchrist, Oliver Pain, Petroula Proitsi, Cathryn M Lewis

**Author notes:** **Corresponding author:** Dr Julian Mutz; Social, Genetic and Developmental Psychiatry Centre, Institute of Psychiatry, Psychology & Neuroscience, King’s College London, Memory Lane, London SE5 8AF, United Kingdom.

## Abstract

**Background:** Identifying individuals at risk of dementia is essential for prevention and targeted disease-modifying strategies. We investigated whether metabolomic ageing in mid-life is associated with incident dementia and dementia age of onset. We also explored joint associations and interactions with genetic risk, including *APOE* genotype and dementia polygenic scores.

**Methods:** The UK Biobank is a community-based observational study of middle-aged and older adults. The Nightingale Health platform was used to quantify plasma metabolites at baseline. Metabolomic age (MileAge) delta represents the difference between metabolite-predicted and chronological age. Dementia cases were identified from primary care, hospital inpatient, death registry and self-reported physician diagnosis data. Associations between MileAge delta and incident dementia were estimated using Cox proportional hazards models.

**Results:** Amongst 223,496 participants, 3976 developed dementia. A metabolite-predicted age exceeding chronological age was associated with higher hazards of all-cause, unspecified and vascular dementia (HR = 1.61, 95% CI 1.28-2.02, *p* = 0.001). A higher MileAge delta was also associated with an earlier dementia onset. Key contributors to the MileAge clock included lipids, lipoproteins and amino acids, which were also associated with dementia. Genetic risk and MileAge delta were jointly associated with incident dementia. For example, individuals with a MileAge exceeding chronological age and two copies of the *APOE* ε4 allele had 10.27-fold higher hazards of all-cause dementia (95% CI 7.93-13.30, *p* < 0.001).

**Conclusions:** Metabolomic ageing in mid-life was associated with incident vascular, unspecified and all-cause dementia, as well as an earlier onset, including of Alzheimer’s disease. Genetic risk and MileAge delta were jointly associated with incident dementia, likely representing independent pathways.

## Introduction

Dementia currently affects an estimated 982,000 individuals in the UK, with projections indicating a rise to 1.4 million by 2040 (Alzheimer’s Research UK, 2024; Wittenberg, Hu, Barraza-Araiza, & Rehill, 2019). In 2023, dementia and Alzheimer’s disease were the leading causes of death in England and Wales, accounting for 11.6% of all registered deaths (Office for National Statistics, 2024). Although chronological age remains the strongest risk factor, dementia is not an inevitable consequence of ageing. Up to 45% of dementia cases could be delayed or prevented through modifying risk factors (Livingston et al., 2024).

Effective risk stratification is essential for targeted prevention and disease-modifying strategies. Despite the long preclinical period of dementia, research has largely focussed on individuals later in life, where opportunities to intervene are limited. Identifying dementia biomarkers in mid-life could help determine risk earlier, providing greater scope for intervention.

Advances in high-throughput molecular technologies have made it feasible to quantify hundreds or thousands of biological markers–including metabolites–at scale (Rutledge, Oh, & Wyss-Coray, 2022). The pathophysiology of most age-related diseases involves changes in the metabolome (Pietzner et al., 2021). Metabolomic irregularities include, for example, dysregulated lipid levels and lipoprotein concentrations (Panyard, Yu, & Snyder, 2022). Given that metabolites can be measured in minimally invasive blood samples, metabolomics holds promise for early disease detection.

Emerging evidence suggests that metabolomic profiles can predict dementia risk. For example, in the Whitehall II study, an elastic net model based on serum metabolites modestly improved prediction of dementia across 20 years (Machado-Fragua et al., 2022). Five metabolites were nominally associated with incident dementia after adjusting for chronological age and sociodemographic factors. Only glucose remained statistically significant after Bonferroni correction, potentially due to its links with neurotoxicity, hyperglycaemia, insulin resistance and vascular injury. In the UK Biobank, metabolomic profiles derived from plasma predicted incident diseases (Buergel et al., 2022). The odds of dementia were 6.39-fold higher in the top 10% of the distribution compared to the bottom 10%. Metabolomic scores developed in the UK Biobank also demonstrated cross-cohort replicability at predicting four-year incidence of Alzheimer’s disease and vascular/other dementia (Barrett et al., 2024).

While metabolomic profiles aid risk stratification, biological ageing clocks are conceptually easier to understand, being expressed in unit of years. Ageing clocks involve using machine learning to identify patterns in biological data to predict a person’s age (Rutledge et al., 2022) and have been developed using diverse molecular markers, such as DNA methylation, protein abundance or metabolite concentrations. We have previously shown that a metabolite-predicted age exceeding chronological age was associated with mortality and diverse health and ageing markers (Mutz, Iniesta, & Lewis, 2024). Ageing clocks also offer certain advantages over clinical predictors such as frailty (Petermann-Rocha et al., 2020) because they can be applied to individuals free of diseases, impairments and functional deficits.

To investigate whether metabolomic ageing predicts dementia, we estimated associations between metabolomic age (MileAge) delta and incident Alzheimer’s disease, vascular dementia, dementia in other diseases, unspecified dementia and all-cause dementia, as well as dementia age of onset. To identify specific metabolites that contributed to the MileAge clock and were associated with dementia, we performed metabolome-wide association analyses. Finally, we examined joint associations and interactions of MileAge delta and genetic risk factors with incident dementia, including apolipoprotein E (*APOE*) genotype and ancestry-standardised dementia polygenic scores.

## Methods

### Study population

UK Biobank recruited over 500,000 adults aged 37–73 years (Bycroft et al., 2018), registered with the UK National Health Service (NHS) and residing within 25-miles of one of 22 assessment centres. At baseline (2006–2010), participants provided sociodemographic, behavioural and medical information, underwent physical examinations and gave blood and urine samples. Hospital inpatient records are available for most participants; primary care records for ∼230,000. Some participants attended follow-up assessments. Table S1 lists all data fields used.

### Metabolomic ageing (MileAge) clock

Nuclear magnetic resonance (NMR) metabolomic biomarkers were quantified in non-fasting plasma samples. The Nightingale Health platform measured 168 metabolites in absolute concentration units using a standardised protocol (Würtz et al., 2017). Technical variation was removed using the ‘ukbnmr’ R package (algorithm v2) (Ritchie et al., 2023). In a previous study (Mutz et al., 2024), we developed a metabolomic clock using Cubist rule-based regression (Kuhn & Johnson, 2013; Quinlan, 1992). Individual-level age predictions were aggregated from the outer nested cross-validation loop test sets to avoid overfitting. Metabolomic age (MileAge) delta was defined as the difference between metabolite-predicted and chronological age.

### Dementia diagnosis

Incident dementia was defined using ICD-10 codes for Alzheimer’s disease (F00, G30), vascular dementia (F01), dementia in other diseases (F02) and unspecified dementia (F03); all-cause dementia included any of these. First occurrence dates were ascertained from primary care, hospital inpatient, death registry and self-reported physician diagnosis data. ICD-10 codes were extracted from hospital and death records. ICD-9 codes from hospital records and Read v2 or CTV3 codes from primary care were extracted if they could be mapped to ICD-10. Administrative, procedure and medication CTV3 codes were not considered. Self-reported physician diagnosis data were mapped to ICD-10, with occurrence dates interpolated to mid-year. Follow-up spanned baseline to dementia diagnosis, loss to follow-up, death or censoring. Region-specific censoring dates based on hospital records were 31 October 2022 (England), 31 August 2022 (Scotland) and 31 May 2022 (Wales).

### Dementia age of onset

We derived approximate birthdates from birth year and month, assigning random days within each month, accounting for the number of days per month and leap years. Age of onset was calculated as days from birth to first occurrence divided by 365.25.

### *APOE* genotype

Apolipoprotein E (*APOE*) genotype was derived from imputed genotype data using single nucleotide polymorphism (SNPs) rs7412 and rs429358 within the *APOE* gene on chromosome 19, via PLINK v2.0 (Chang et al., 2015). SNPs were combined to infer *APOE* alleles (ε1, ε2, ε3 and ε4) (Lumsden, Mulugeta, Zhou, & Hyppönen, 2020). Ambiguous genotypes (ε1ε3 and ε2ε4) were coded as ε2ε4; rare genotypes (ε1ε2 and ε1ε4) were excluded. We defined four risk categories: low (ε2ε2 and ε2ε3), average (ε3ε3), moderate (ε2ε4 and ε4ε3) and high (ε4ε4), with ε3ε3 serving as reference.

### Dementia polygenic scores

Genotype quality control steps are reported in Supplementary Methods; genome-wide association study (GWAS) summary statistics in Table S2. Polygenic scores (PRS) were calculated using GenoPred (Pain, Al-Chalabi, & Lewis, 2024). A combined 1000 Genomes Project phase 3 (Auton et al., 2015) and Human Genome Diversity Project (Bergström et al., 2020) reference panel, restricted to 1,204,449 HapMap3 variants, was used for linkage disequilibrium estimation. MegaPRS (LDAK v5.1) was used for polygenic scoring, applying the BLD-LDAK heritability model where SNPs are weighted based on allele frequency, linkage disequilibrium and functional annotations (Zhang, Privé, Vilhjálmsson, & Speed, 2021). MegaPRS models for each dementia outcome were selected using pseudo-validation. PRS were calculated within assigned ancestry groups (matching individuals to populations in the *N* = 3313 reference data with probability ≥ 0.95) and scaled to units of standard deviation from the matched reference population mean (Pain et al., 2021). PRS subgroups were defined on this scale.

### Covariates

A directed acyclic graph (DAG) was used to visualise relationships between the exposure (MileAge delta), outcome (dementia) and select confounders (chronological age, sex, highest educational/professional qualification, cohabitation with spouse/partner, annual gross household income, Townsend deprivation index, fasting time and *APOE* genotype) (Figure S1). Analyses of *APOE* genotype and PRS also included genotype batch number, assessment centre and the first six population principal components (derived within UK Biobank) to account for population stratification (Coleman et al., 2020).

### Exclusion criteria

We excluded women with possible pregnancy (due to altered metabolite profiles), individuals with discordant genetic and self-reported sex (to mitigate potential data quality issues) and missing or outlier metabolite values 4× the interquartile range (IQR) from the median. Individuals with a dementia onset preceding baseline (i.e., prevalent cases) and those without an event date, with a date prior to, matching or in the same calendar year as their birth date or with a future date beyond the follow-up period were also excluded.

### Statistical analyses

Analyses were performed in R (version 4.3.0). Sample characteristics were summarised using means and standard deviations or counts and percentages. We estimated hazard ratios (HRs) and 95% confidence intervals using Cox proportional hazards models to investigate associations between MileAge delta and incident dementia (separate models for Alzheimer’s disease, vascular dementia, dementia in other diseases, unspecified dementia and all-cause dementia). For the primary analysis, MileAge delta subgroups were defined by standard deviation from the mean. Individuals at least one standard deviation below the mean (i.e., with a younger metabolite-predicted age) served as the reference. Time (in days) since baseline was the underlying time axis. Model 1 was adjusted for chronological age and sex; Model 2 additionally included education, cohabitation, income, deprivation and fasting time; Model 2B was further adjusted for *APOE* genotype.

To explore non-linear relationships between MileAge delta and dementia, we fitted Cox proportional hazard models with penalised splines adjusted for Model 2 covariates. Associations between MileAge delta and dementia age of onset were estimated using ordinary least squares regression, with the covariate adjustment from our primary analysis. To identify metabolites that were important contributors to the MileAge clock and associated with incident dementia, we performed metabolome-wide association analyses using Cox proportional hazard models (adjusted for Model 2 covariates to reduce the possibility of confounding). We visualised the hazard ratios from these analyses alongside variable importance (VIP) scores previously calculated for MileAge (Mutz et al., 2024).

We also investigated joint associations and interactions with genetic risk. First, we estimated differences in MileAge delta between *APOE* genotypes and associations with dementia polygenic scores (across and within ancestry groups), using ordinary least squares regression. We then estimated associations between MileAge delta and incident dementia by *APOE* genotype risk category, using Cox proportional hazard models adjusted for age and sex (Model 1) and for all covariates (Model 2). We performed similar analyses for PRS groups, classified into low (≤ 1 SD below the mean), middle and high (≥ 1 SD above the mean) risk categories. Individuals with a MileAge delta in the middle of the distribution in the average *APOE* risk category or middle PRS group served as reference. Given that few incident dementia cases were observed in non-Europeans (Table S3), these analyses were performed only across ancestry groups.

### Sensitivity analyses

We performed several analyses to assess the robustness of our findings. We excluded self-reported physician diagnosis data, as the first occurrence date was approximated and these data are prone to recall error. Individuals with another data source with a matching or subsequent event date were retained. To reduce the possibility of reverse causality, we excluded individuals with incident dementia within two years since baseline and those with less than two years of follow up. We also restricted analyses to those aged 60 years or older at baseline, representing those at risk of dementia. Given that only 45% of participants had linked primary care records, we performed a sensitivity analysis within this subset. Dementia may be first or exclusively identified in primary care, potentially resulting in misclassification in the comparison group or inaccurate first occurrence dates. Region and data-provider specific censoring dates were 31 August 2017 (Wales), 31 May 2017 (England, Vision), 31 March 2017 (Scotland) and 31 May 2016 (England, TPP). To address diagnostic uncertainty, we excluded individuals with multiple diagnostic labels. Since certain treatments could impact observed associations, we also excluded individuals using lipid-modifying treatments at baseline (Table S4).

## Results

### Sample characteristics

Among 274,315 participants with metabolomics data, 247,861 had complete data (Figure 1A). Excluding individuals with outlier metabolite values, discordant self-reported and genetic sex, possible pregnancy, prevalent dementia or missing *APOE* genotype, the sample comprised 223,496 participants (54% female; mean age = 56.97 years, SD = 8.10) (Table 1).

**Figure 1.**
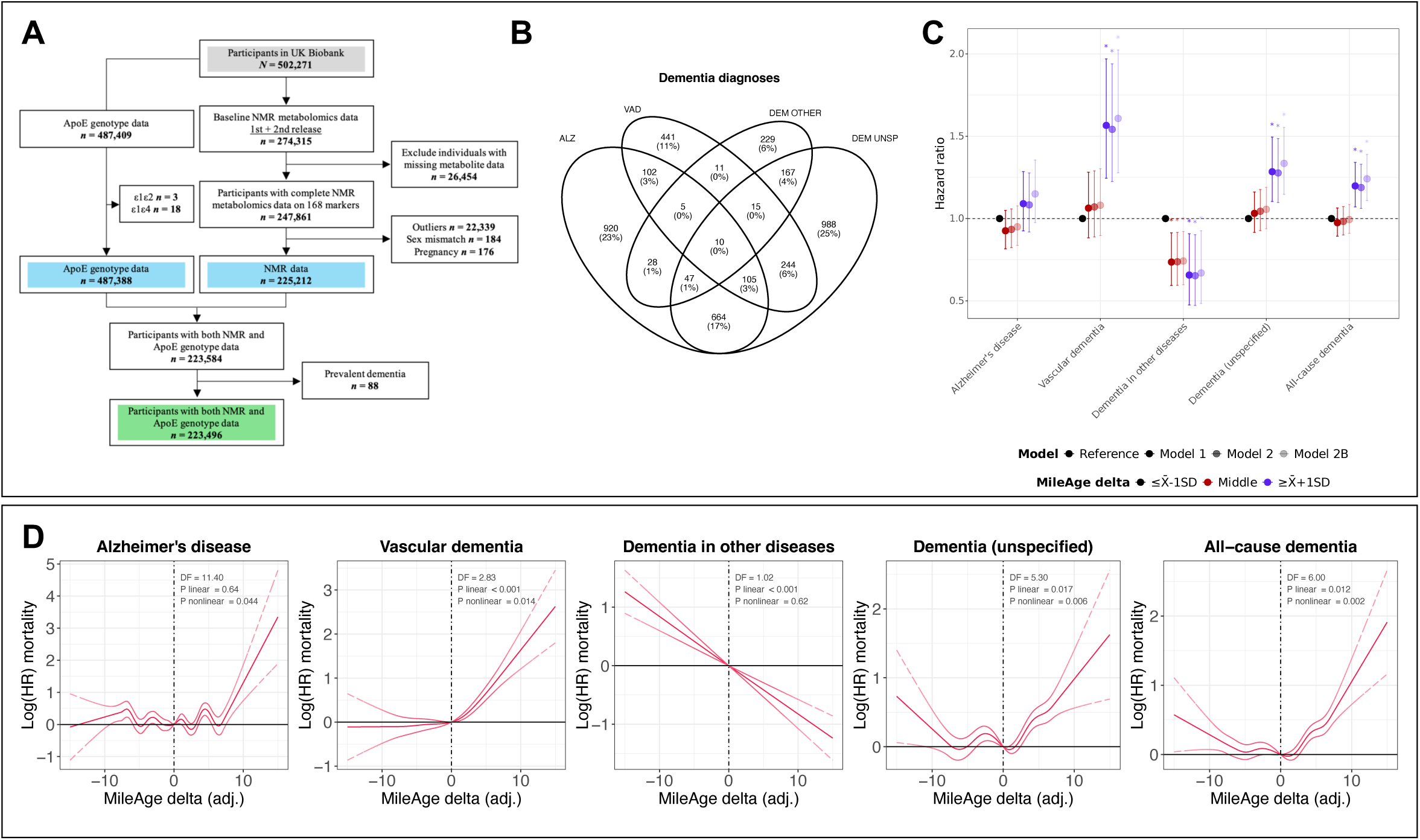
Sample overview and incident dementia. **(A)** Study sample flowchart. Outliers were defined as metabolite values 4×IQR above or below the median. NMR = nuclear magnetic resonance; IQR = interquartile range. **(B)** Venn diagram of incident dementia cases. ALZ = Alzheimer’s disease; VAD = vascular dementia; DEM OTHER = dementia in other diseases; DEM UNSP = unspecified dementia. **(C)** Hazard ratios (HR) and 95% confidence intervals from Cox proportional hazards models for incident dementia by MileAge delta. Time since the baseline assessment (in days) was used as the underlying time axis. Reference group: individuals with a MileAge delta smaller than one standard deviation below the mean. Model 1–adjusted for age and sex; Model 2–adjusted for age, sex, highest educational/professional qualification, cohabitation with spouse/partner, annual gross household income, Townsend deprivation index and fasting time; Model 2B–additionally adjusted for apolipoprotein E (*APOE*) genotype. Asterisks indicate statistical significance after correcting *p*-values for multiple testing using the Benjamini–Hochberg procedure. **(D)** Log(HRs) and 95% confidence intervals from Cox proportional hazards models for incident dementia, adjusted for Model 2 covariates. Time since the baseline assessment (in days) was used as the underlying time axis. A penalised spline function was applied, with degrees of freedom (DF) selected based on Akaike Information Criterion. Vertical lines indicate the values of the distribution closest to zero (0.00001192219), representing the reference for interpreting the estimates shown. **(C** and **D)** *N* = 1881 (Alzheimer’s disease); *N* = 933 (vascular dementia); *N* = 512 (dementia in other diseases); *N* = 2240 (unspecified dementia); *N* = 3976 (all-cause dementia).

**Table 1.** Sample characteristics.

|  | Analytical sample |  |  |
| --- | --- | --- | --- |
|  | No dementia<br>(N=219,520) | Incident dementia<br>(N=3976) | Full sample<br>(N=223,496) |
| MileAge delta, mean (SD) | 0.00 (3.77) | -0.06 (3.73) | 0.00 (3.77) |
| Age, mean (SD) | 56.82 (8.08) | 65.04 (4.32) | 56.97 (8.10) |
| Sex |  |  |  |
| Female | 118716 (54.1%) | 1885 (47.4%) | 120601 (54.0%) |
| Male | 100804 (45.9%) | 2091 (52.6%) | 102895 (46.0%) |
| Ethnicity |  |  |  |
| White | 207991 (94.7%) | 3831 (96.4%) | 211822 (94.8%) |
| Mixed | 1230 (0.6%) | 15 (0.4%) | 1245 (0.6%) |
| Black | 3103 (1.4%) | 45 (1.1%) | 3148 (1.4%) |
| Asian | 3773 (1.7%) | 43 (1.1%) | 3816 (1.7%) |
| Chinese | 631 (0.3%) | 2 (0.1%) | 633 (0.3%) |
| Other | 1828 (0.8%) | 16 (0.4%) | 1844 (0.8%) |
| Missing<sup>1</sup> | 964 (0.4%) | 24 (0.6%) | 988 (0.4%) |
| Highest qualification |  |  |  |
| None | 37034 (16.9%) | 1405 (35.3%) | 38439 (17.2%) |
| O levels/GCSEs/CSEs | 59408 (27.1%) | 821 (20.6%) | 60229 (26.9%) |
| A levels/NVQ/HND/HNC<sup>2</sup> | 50241 (22.9%) | 886 (22.3%) | 51127 (22.9%) |
| Degree | 70364 (32.1%) | 777 (19.5%) | 71141 (31.8%) |
| Missing<sup>1</sup> | 2473 (1.1%) | 87 (2.2%) | 2560 (1.1%) |
| Household income<sup>3</sup> |  |  |  |
| Very low | 42134 (19.2%) | 1421 (35.7%) | 43555 (19.5%) |
| Low | 48375 (22.0%) | 919 (23.1%) | 49294 (22.1%) |
| Medium | 49531 (22.6%) | 462 (11.6%) | 49993 (22.4%) |
| High | 38203 (17.4%) | 209 (5.3%) | 38412 (17.2%) |
| Very high | 9682 (4.4%) | 42 (1.1%) | 9724 (4.4%) |
| Missing<sup>1</sup> | 31595 (14.4%) | 923 (23.2%) | 32518 (14.5%) |
| Townsend deprivation |  |  |  |
| Q1 | 45557 (20.8%) | 811 (20.4%) | 46368 (20.7%) |
| Q2 | 45405 (20.7%) | 782 (19.7%) | 46187 (20.7%) |
| Q3 | 44080 (20.1%) | 762 (19.2%) | 44842 (20.1%) |
| Q4 | 42894 (19.5%) | 748 (18.8%) | 43642 (19.5%) |
| Q5 | 41315 (18.8%) | 869 (21.9%) | 42184 (18.9%) |
| Missing<sup>1</sup> | 269 (0.1%) | 4 (0.1%) | 273 (0.1%) |
| Cohabitation |  |  |  |
| With partner | 160618 (73.2%) | 2752 (69.2%) | 163370 (73.1%) |
| Single | 18011 (8.2%) | 192 (4.8%) | 18203 (8.1%) |
| Missing<sup>1</sup> | 40891 (18.6%) | 1032 (26.0%) | 41923 (18.8%) |
| Fasting time |  |  |  |
| Less than 8 hours | 210941 (96.1%) | 3833 (96.4%) | 214774 (96.1%) |
| At least 8 hours | 8575 (3.9%) | 143 (3.6%) | 8718 (3.9%) |
| Missing<sup>1</sup> | 4 (0.0%) | 0 (0.0%) | 4 (0.0%) |
| Lipid-modifying treatment |  |  |  |
| Yes | 175138 (79.8%) | 2247 (56.5%) | 177385 (79.4%) |
| No | 43386 (19.8%) | 1693 (42.6%) | 45079 (20.2%) |
| Missing<sup>1</sup> | 996 (0.5%) | 36 (0.9%) | 1032 (0.5%) |
| APOE genotype |  |  |  |
| ε3ε3 | 129685 (59.1%) | 1551 (39.0%) | 131236 (58.7%) |
| ε2ε2 | 1333 (0.6%) | 10 (0.3%) | 1343 (0.6%) |
| ε2ε3 | 26920 (12.3%) | 265 (6.7%) | 27185 (12.2%) |
| ε2ε4 | 5531 (2.5%) | 124 (3.1%) | 5655 (2.5%) |
| ε4ε3 | 51128 (23.3%) | 1591 (40.0%) | 52719 (23.6%) |
| ε4ε4 | 4923 (2.2%) | 435 (10.9%) | 5358 (2.4%) |

Note: SD = standard deviation. GCSEs = general certificate of secondary education; CSE = certificate of secondary education; NVQ = national vocational qualification; HND = higher national diploma; HNC = higher national certificate. <sup>1</sup> Missing data may also include “do not know” or “prefer not to answer”. <sup>2</sup> Also includes ‘other professional qualifications’. <sup>3</sup> Annual household income groups: very low (<£18,000), low (£18,000–£30,999), middle (£31,000–£51,999), high (£52,000–£100,000) and very high (>£100,000).

The median follow-up of censored individuals was 13.7 years (IQR = 1.4), with up to 2,971,915 person-years. There were 3976 incident all-cause dementia cases: 1881 with Alzheimer’s disease, 933 with vascular dementia, 512 with dementia in other diseases and 2240 with unspecified dementia. Most were identified from hospital records (Table S5); 35% (*N* = 1404) of participants carried multiple diagnostic labels (Figure 1B).

### MileAge delta and incident dementia

A metabolite-predicted age exceeding chronological age was associated with higher hazards of vascular, unspecified and all-cause dementia (Figure 1C). The age and sex-adjusted hazard ratio (HR) for all-cause dementia was 1.20 (95% CI 1.07-1.34, *p* = 0.006). Further adjustment for education, cohabitation status, income, deprivation, fasting time and *APOE* genotype had little impact on these estimates (Table 2). MileAge delta showed the strongest association with vascular dementia (HR = 1.61, 95% CI 1.28-2.02, *p* = 0.001 after full adjustment). No statistically significant associations were identified for Alzheimer’s disease (*p*-values 0.119– 0.477). MileAge delta was associated with a lower hazard of dementia in other diseases (HR = 0.67, 95% CI 0.48-0.93, *p* = 0.031). There was evidence of non-linear associations between MileAge delta and dementia, except dementia in other diseases (Figure 1D).

**Table 2.** MileAge delta and incident dementia.

|  |  |  | Model 1 |  |  |  | Model 2 |  |  |  | Model 2B |  |  |  |
| --- | --- | --- | --- | --- | --- | --- | --- | --- | --- | --- | --- | --- | --- | --- |
| Level | $N_{\text{total}}$ | $N_{\text{incident}}$ | HR | 95% CI | | $p$ | HR | 95% CI | | $p$ | HR | 95% CI | | $p$ |
| Alzheimer's disease |  |  |  |  |  |  |  |  |  |  |  |  |  |  |
| $\leq \bar{X}$-1SD | 35656 | 301 | Reference | | | | | | | | | | | |
| Middle | 150745 | 1306 | 0.93 | 0.82 | 1.05 | 0.406 | 0.92 | 0.80 | 1.07 | 0.281 | 0.94 | 0.81 | 1.08 | 0.373 |
| $\geq \bar{X}$+1SD | 35000 | 274 | 1.09 | 0.93 | 1.29 | 0.477 | 1.10 | 0.90 | 1.33 | 0.355 | 1.17 | 0.96 | 1.42 | 0.119 |
| Vascular dementia |  |  |  |  |  |  |  |  |  |  |  |  |  |  |
| $\leq \bar{X}$-1SD | 35490 | 135 | Reference | | | | | | | | | | | |
| Middle | 150075 | 636 | 1.06 | 0.88 | 1.28 | 0.596 | 1.07 | 0.89 | 1.29 | 0.570 | 1.08 | 0.90 | 1.30 | 0.551 |
| $\geq \bar{X}$+1SD | 34888 | 162 | 1.57 | 1.24 | 1.97 | 0.001 | 1.54 | 1.23 | 1.94 | 0.001 | 1.61 | 1.28 | 2.02 | 0.001 |
| Dementia in other diseases |  |  |  |  |  |  |  |  |  |  |  |  |  |  |
| $\leq \bar{X}$-1SD | 35464 | 109 | Reference | | | | | | | | | | | |
| Middle | 149786 | 347 | 0.74 | 0.59 | 0.91 | 0.016 | 0.74 | 0.59 | 0.92 | 0.016 | 0.74 | 0.60 | 0.92 | 0.017 |
| $\geq \bar{X}$+1SD | 34782 | 56 | 0.66 | 0.48 | 0.91 | 0.023 | 0.65 | 0.47 | 0.90 | 0.023 | 0.67 | 0.48 | 0.93 | 0.031 |
| Dementia (unspecified) |  |  |  |  |  |  |  |  |  |  |  |  |  |  |
| $\leq \bar{X}$-1SD | 35686 | 331 | Reference | | | | | | | | | | | |
| Middle | 151002 | 1563 | 1.03 | 0.92 | 1.16 | 0.652 | 1.04 | 0.93 | 1.18 | 0.570 | 1.06 | 0.94 | 1.19 | 0.526 |
| $\geq \bar{X}$+1SD | 35072 | 346 | 1.28 | 1.10 | 1.49 | 0.006 | 1.28 | 1.10 | 1.49 | 0.006 | 1.34 | 1.15 | 1.55 | 0.001 |
| All-cause dementia |  |  |  |  |  |  |  |  |  |  |  |  |  |  |
| $\leq \bar{X}$-1SD | 35973 | 618 | Reference | | | | | | | | | | | |
| Middle | 152192 | 2753 | 0.98 | 0.89 | 1.06 | 0.636 | 0.98 | 0.90 | 1.07 | 0.722 | 0.99 | 0.91 | 1.09 | 0.894 |
| $\geq \bar{X}$+1SD | 35331 | 605 | 1.20 | 1.07 | 1.34 | 0.006 | 1.19 | 1.06 | 1.33 | 0.009 | 1.24 | 1.11 | 1.39 | 0.001 |

*Note:* HR = hazard ratio; CI = confidence interval; SD = standard deviation. Time since the baseline assessment (in days) was used as the underlying time axis. Model 1—adjusted for age and sex; Model 2—adjusted for age, sex, highest educational/professional qualification, cohabitation with spouse/partner, annual gross household income, Townsend deprivation index and fasting time; Model 2B—additionally adjusted for apolipoprotein E (*APOE*) genotype.

#### Sensitivity analyses

Excluding self-reported physician diagnoses (*N* = 16), those with dementia within two years of baseline (*N* = 46) and those with less than two years of follow-up (*N* = 1043) had negligible impact (Figures S2-S3; Table S6-S7). Estimates were slightly attenuated when restricting to individuals aged ≥ 60 years at baseline (*N* = 96,243; Figure S4; Table S8). In those with linked primary care data (*N* = 101,952; Figure S5), the median follow-up was 7.43 years (IQR = 1.5). MileAge delta remained nominally associated with vascular dementia (HR = 1.59, 95% CI 1.11-2.29, *p* = 0.135; Figure S6) and with unspecified dementia only after full adjustment (Table S9). Excluding individuals with multiple diagnoses (*N* = 1398) yielded attenuated associations for vascular dementia and dementia in other diseases, which did not reach statistical significance (Figure S7; Table S10). Excluding individuals on lipid-modifying treatments (*N* = 45,079) attenuated most estimates and associations were not statistically significant after multiple testing correction (Figure S8; Table S11).

### MileAge delta and dementia age of onset

Mean age of onset ranged from 74.24 years (SD = 5.20) for dementia in other diseases to 75.57 years (SD = 5.06) for Alzheimer’s disease (Figure 2A; Table S12). Higher MileAge delta was associated with an earlier onset of Alzheimer’s disease (*β =* −0.09, 95% CI −0.15 to −0.02, *p* = 0.009), vascular dementia (*β* = −0.16, 95% CI −0.24 to −0.07, *p* < 0.001), unspecified dementia (*β* = −0.11, 95% CI −0.17 to −0.05, *p* < 0.001) and all-cause dementia (*β* = −0.12, 95% CI −0.16 to −0.07, *p* < 0.001) (Figures 2B and 2C). Covariate adjustment did not materially impact these associations (Table 3).

**Figure 2.**
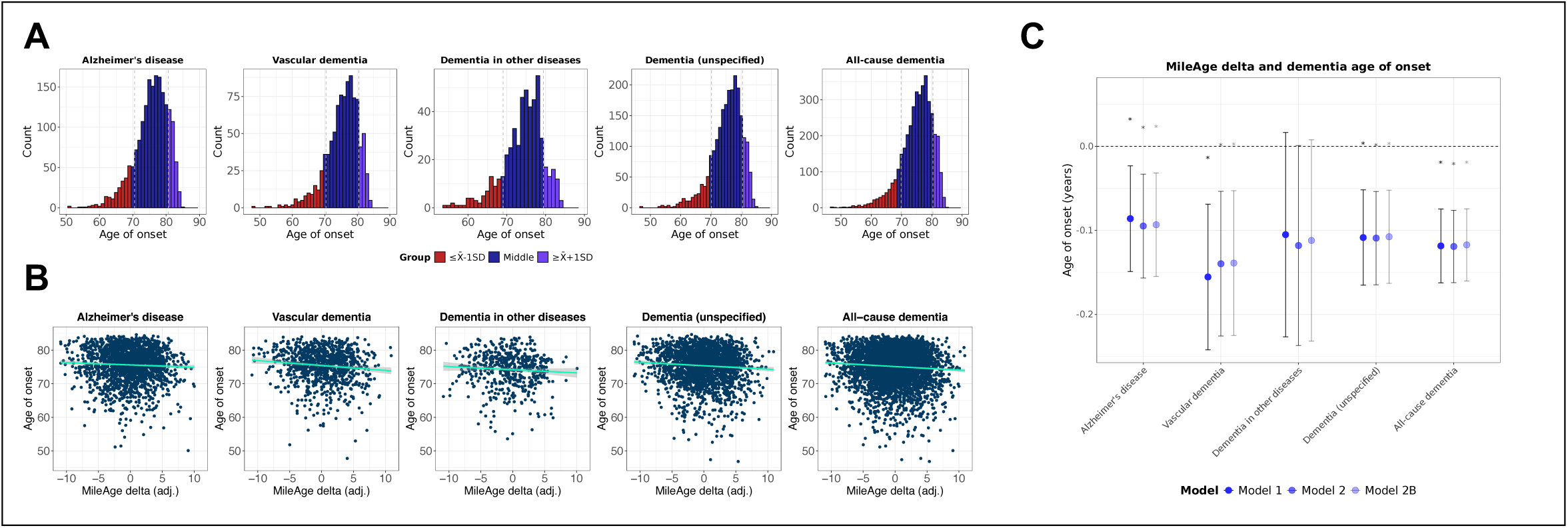
Dementia age of onset. **(A)** Histograms showing the distribution of dementia age of onset. **(B)** Scatter plots showing the association between MileAge delta and dementia age of onset. **(C)** Associations between MileAge delta and dementia age of onset, with betas and 95% confidence intervals estimated using linear regression models. Model 1–adjusted for sex; Model 2–adjusted for sex, highest educational/professional qualification, cohabitation with spouse/partner, annual gross household income, Townsend deprivation index and fasting time; Model 2B–additionally adjusted for apolipoprotein E (*APOE*) genotype. Asterisks indicate statistical significance after correcting *p*-values for multiple testing using the Benjamini–Hochberg procedure. **(A** to **C)** *N* = 1881 (Alzheimer’s disease); *N* = 933 (vascular dementia); *N* = 512 (dementia in other diseases); *N* = 2240 (unspecified dementia); *N* = 3976 (all-cause dementia).

**Table 3.**
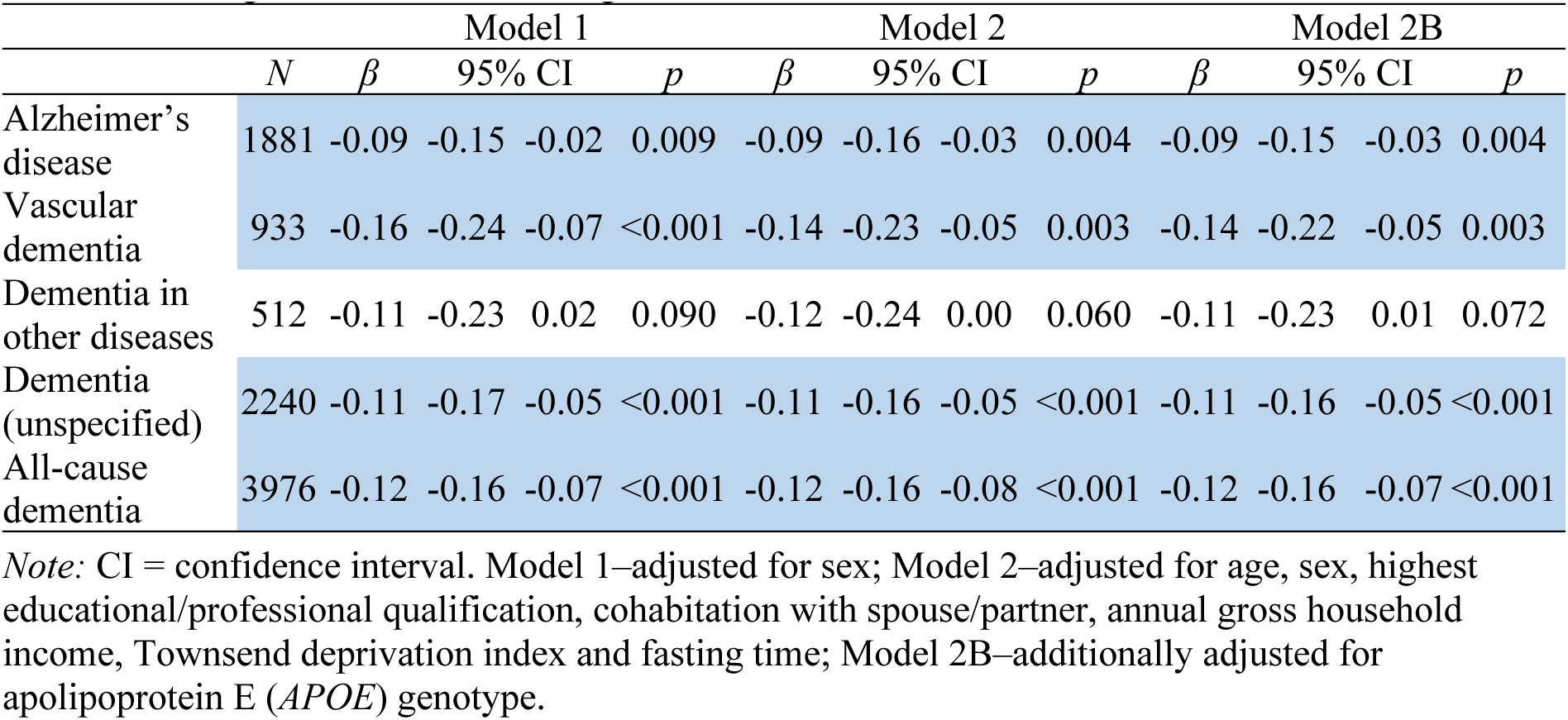
MileAge delta and dementia age of onset.

### Metabolome-wide association analyses

Amongst 69 metabolites nominally associated with Alzheimer’s disease (*p* < 0.05), eight remained statistically significant after Bonferroni correction: leucine, valine, total branched-chain amino acids and certain lipoprotein measures—all associated with a lower hazard of Alzheimer’s disease (Figure 3A). Of the 101 metabolites nominally associated with vascular dementia, 72 survived multiple testing correction. GlycA was the only metabolite associated with a higher hazard of vascular dementia (HR = 1.14, 95% CI 1.07-1.22, *p* < 0.001), while lipids and lipoproteins were associated with a lower hazard (estimates ranging from HR = 0.79 for linoleic acid to HR = 0.88 for very low-density lipoprotein phospholipids). None of the 54 metabolites associated with dementia in other diseases after Bonferroni correction had a HR > 1; lipids and lipoproteins were associated with a HR < 1 (e.g., polyunsaturated fatty acids: HR = 0.78, 95% CI 0.71-0.86, *p* < 0.001). After multiple testing correction, 92 and 106 metabolites remained statistically significantly for unspecified and all-cause dementia. Glucose-lactate, a composite measure that accounts for anaerobic glycolysis between sample collection and processing, was associated with higher hazards of all-cause dementia (HR = 1.06, 95% CI 1.03-1.09, *p* < 0.001) and nominally associated with unspecified dementia (HR = 1.07, 95% CI 1.03-1.11, *p* < 0.001). Lipids, lipoproteins and amino acids (leucine, valine and total branched-chain amino acids) were associated with lower hazards of unspecified and all-cause dementia (Additional File 1).

**Figure 3.**
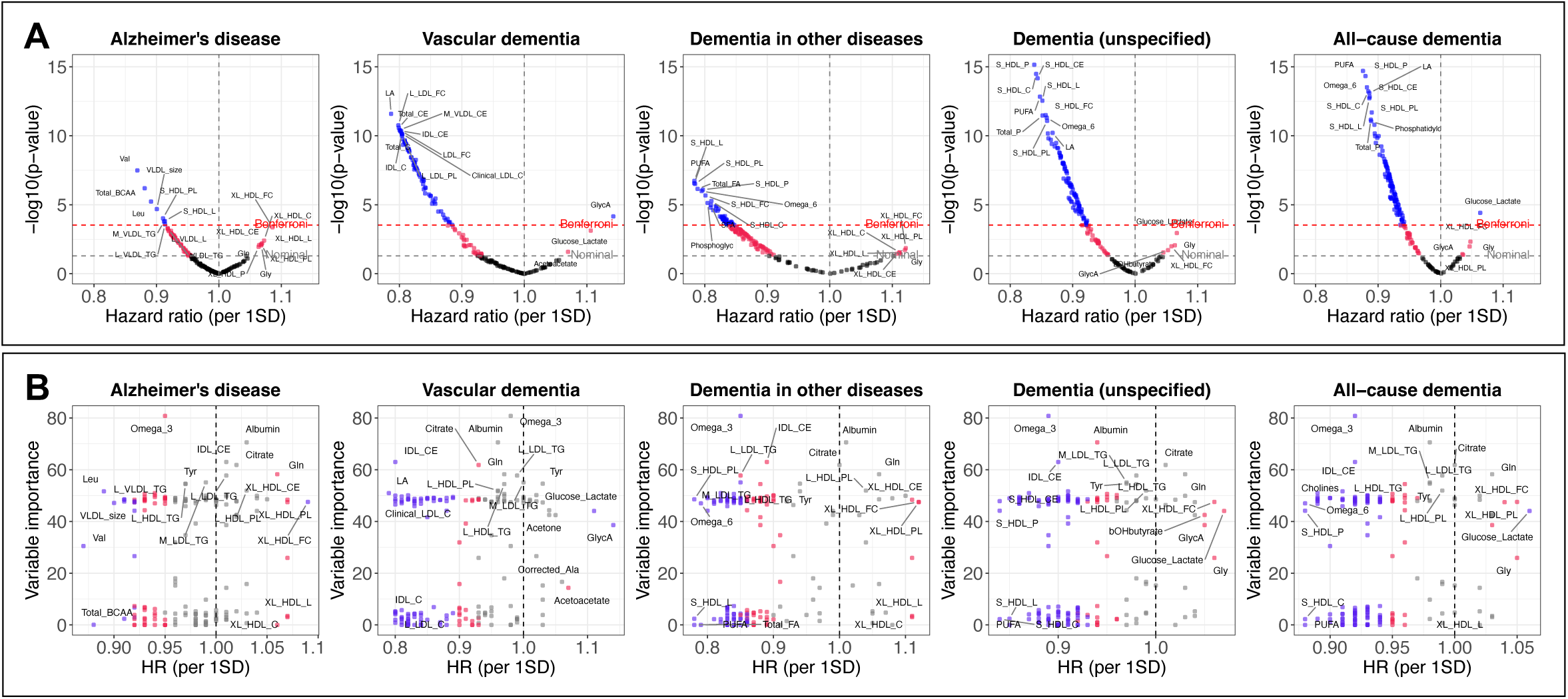
Metabolome-wide associations. **(A)** Volcano plots showing associations between metabolite levels and incident dementia. Metabolites were standardised to have a mean equal to zero and a standard deviation (SD) of one. Hazard ratios (HR) were estimated using Cox proportional hazards models, with days since the baseline assessment as the underlying time axis. Analyses were adjusted for age, sex, highest educational/professional qualification, cohabitation with spouse/partner, annual gross household income, Townsend deprivation index and fasting time. *P*-values were -log10 transformed. **(B)** Scatter plots showing HRs for incident dementia associated with a one standard deviation difference in metabolite levels and MileAge variable importance (VIP) scores. VIPs represent averages across the 10 outer loops of the nested cross-validation and are the same across the five panels. Metabolites with statistically significant associations with incident dementia (based on their HRs) are shown in red and purple, indicating nominal and Bonferroni-corrected significant levels, respectively. **(A** and **B)** *N* = 223,496.

Certain metabolites associated with dementia also strongly impacted the predictive accuracy of MileAge (Figure 3B). Omega 3 had the highest variable importance (VIP) score of 80.8 and inversely associated with Alzheimer’s disease, dementia in other diseases, unspecified dementia and all-cause dementia (at *p* < 0.05/168). Albumin had the second highest VIP score (70.6) but was not associated with dementia after multiple testing correction. Intermediate-density lipoprotein cholesteryl esters (VIP = 63.0) were associated with lower hazards of vascular, unspecified and all-cause dementia (Bonferroni-corrected). However, there was no strong correlation between VIP scores and dementia hazard ratios. For example, polyunsaturated fatty acids were associated with all dementia outcomes but had a VIP score of zero (Additional file 1).

### MileAge delta and dementia polygenic scores

Amongst 219,481 individuals with genotype data, polygenic scores (PRS) for Alzheimer’s disease were associated with a metabolite-predicted age younger than chronological age (*β* = −0.039, 95% CI −0.054 to −0.023, *p* < 0.001) (Figure 4A). The magnitude of this association corresponded to a difference in MileAge delta of about two weeks per one standard deviation difference in PRS (Table S13). In ancestry-specific analyses, PRS for Alzheimer’s disease was associated with a younger metabolite-predicted age in Europeans (*β* = −0.038, 95% CI - 0.053 to −0.022, *p* < 0.001) and Central/South Asians (*β* = −0.174, 95% CI −0.300 to −0.047, *p* = 0.019) and with a nominally older metabolite-predicted age in East Asians (*β* = 0.363, 95% CI 0.096-0.630, *p* = 0.052) (Figure 4B). The non-European estimates corresponded to differences in MileAge delta of about two and four months, respectively, per standard deviation difference in PRS. In Central/South Asians, PRS for unspecified and all-cause dementia were associated with a younger metabolite-predicted age (*β* = −0.230, 95% CI - 0.359 to −0.102, *p* = 0.005 and *β* = −0.219, 95% CI −0.350 to −0.087, *p* = 0.005) (Table S14).

**Figure 4.**
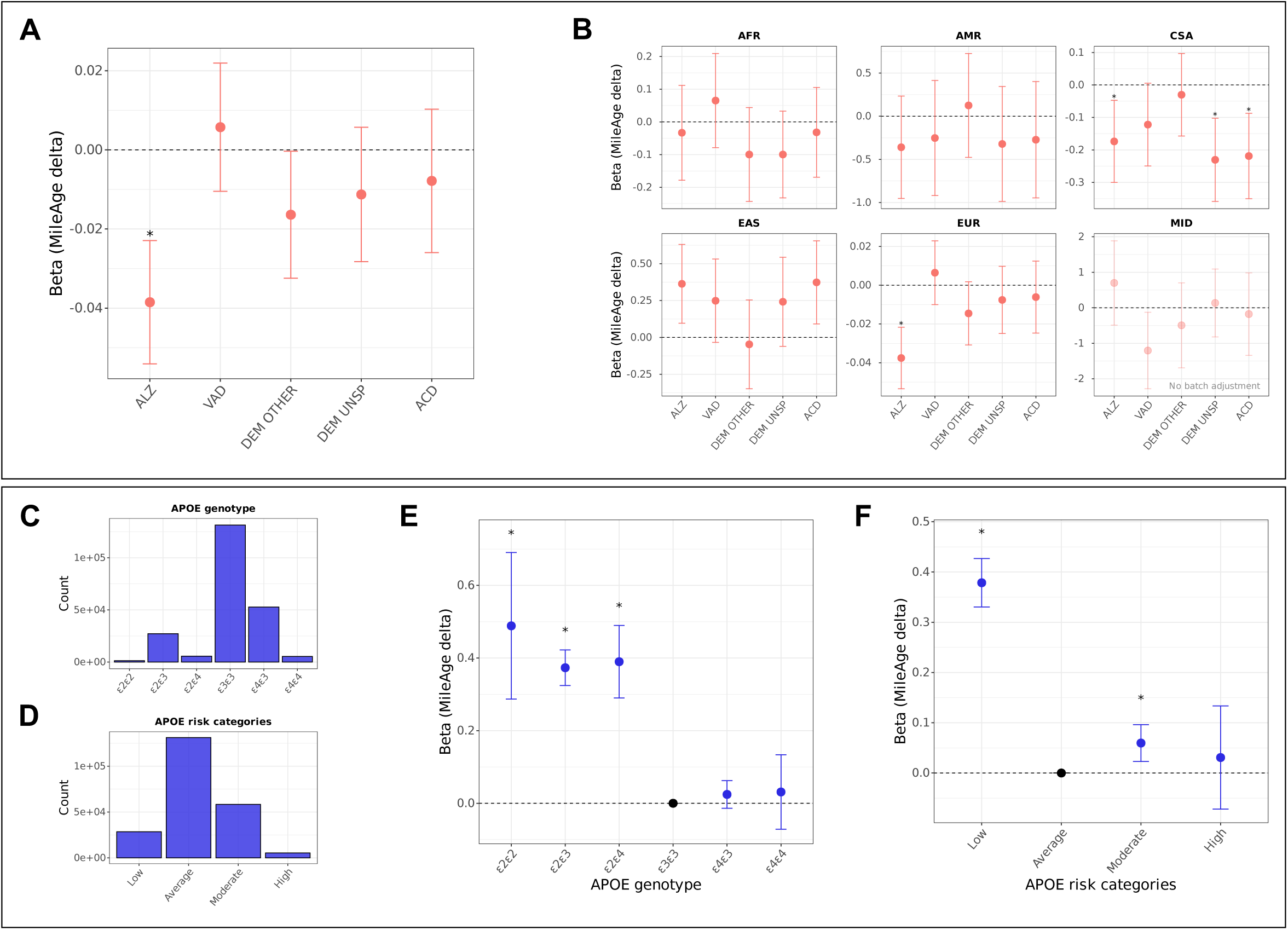
Dementia polygenic scores and *APOE* genotype. **(A)** Associations between dementia polygenic scores and MileAge delta. ALZ = Alzheimer’s disease; VAD = vascular dementia; DEM OTHER = dementia in other diseases; DEM UNSP = unspecified dementia; ACD = all-cause dementia. *N* = 219,481. **(B)** Associations between dementia polygenic scores and MileAge delta by ancestry population. AFR = African; AMR = Admixed American; EAS = East Asian; EUR = European; CSA = Central and South Asian; MID = Middle Eastern. *N* = 3426 (AFR); *N* = 278 (AMR); *N* = 3916 (CSA); *N* = 1012 (EAS); *N* = 210,755 (EUR); *N* = 94 (MID). **(C)** Bar plot showing the distribution of apolipoprotein E (*APOE*) genotypes. **(D)** Bar plot showing the distribution of *APOE* genotypes classified into low (ε2ε2 and ε2ε3), average (ε3ε3), moderate (ε2ε4 and ε4ε3) and high (ε4ε4) risk categories. **(E)** Associations between *APOE* genotypes and MileAge delta. **(F)** Associations between *APOE* genotype risk categories and MileAge delta. **(A**, **B**, **E** and **F)** Estimates shown are ordinary least squares regression beta coefficients and 95% confidence intervals. Models were adjusted for the first six population principal components, genotype batch number, assessment centre, age, sex, highest educational/professional qualification, cohabitation with spouse/partner, annual gross household income, Townsend deprivation index and fasting time. Asterisks indicate statistical significance after correcting *p*-values for multiple testing using the Benjamini–Hochberg procedure. **(C** to **F)** *N* = 223,496.

### MileAge delta and *APOE* genotype

*APOE* genotype and risk category distributions are shown in Figures 4C and 4D. *APOE* genotypes ε2ε2, ε2ε3 and ε2ε4 were associated with a metabolite-predicted age older than chronological age, with betas ranging from 0.373 to 0.489 (Figure 4E; Table S15). Compared to average risk (ε3ε3), low and moderate risk, but not high risk, was associated with a MileAge exceeding chronological age (Figure 4F).

### MileAge delta and incident dementia by *APOE* genotype

Individuals with ε2ε2/ε2ε3 genotypes had lower hazards of dementia (HRs between 0.70 and 0.86), whereas those with at least one ε4 allele had higher hazards of dementia (up to HR = 12.41, 95% CI 11.26-13.68, *p* < 0.001 for Alzheimer’s disease) (Figure S9; Table S16). Individuals in the moderate (ε2ε4 and ε4ε3) and high (ε4ε4) *APOE* risk groups had elevated hazards of dementia (Table S17). Hazards were generally highest in those with a metabolite-predicted age exceeding chronological age and lowest in those with a younger metabolite-predicted age (Figure 5A). For example, the hazard ratios for all-cause dementia were 3.11 (95% CI 2.70-3.57, *p* < 0.001) and 10.27 (95% CI 7.93-13.30, *p* < 0.001) for individuals with a metabolite-predicted age that exceeded chronological age in the moderate and high *APOE* risk groups, respectively.

**Figure 5.**
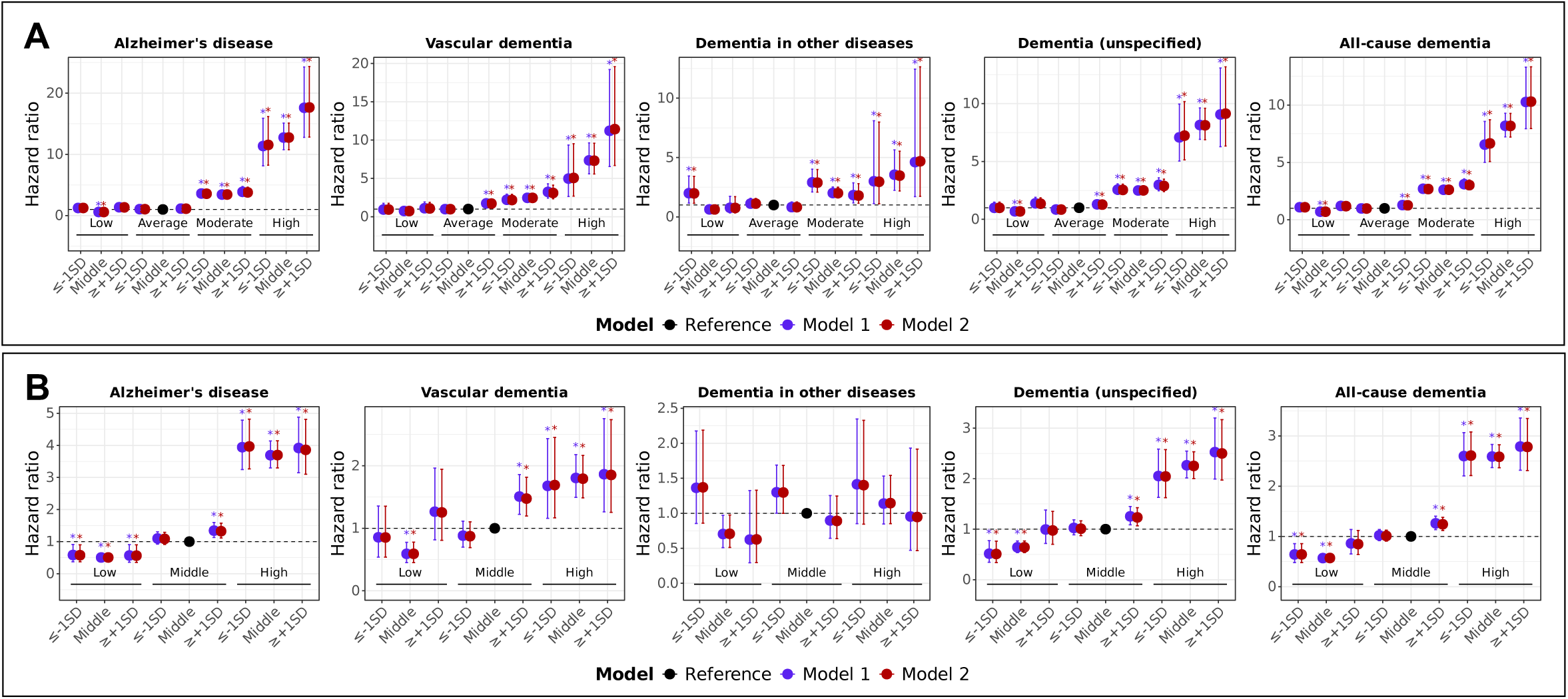
MileAge delta and incident dementia by *APOE* genotype and dementia PRS. **(A)** Hazard ratios and 95% confidence intervals from Cox proportional hazards models for incident dementia by MileAge delta and apolipoprotein E (*APOE*) genotypes. *APOE* genotypes were classified into low (ε2ε2 and ε2ε3), average (ε3ε3), moderate (ε2ε4 and ε4ε3) and high (ε4ε4) risk categories. Time since the baseline assessment (in days) was used as the underlying time axis. Reference group: individuals in the average *APOE* risk category with a MileAge delta in the middle of the distribution. Model 1–adjusted for the first six population principal components, age and sex; Model 2–adjusted for the first six population principal components, age, sex, highest educational/professional qualification, cohabitation with spouse/partner and annual gross household income. No adjustment was made for genotype batch number, assessment centre, Townsend deprivation index and fasting time due to sparse data. SD = standard deviation. **(B)** Hazard ratios and 95% confidence intervals from Cox proportional hazards models for incident dementia by MileAge delta and dementia polygenic scores (PRS). PRS were classified into low (≤ 1 SD below the mean), middle and high (≥ 1 SD above the mean) risk categories. Time since the baseline assessment (in days) was used as the underlying time axis. Reference group: individuals with a PRS and MileAge delta in the middle of the distribution. Model 1–adjusted for the first six population principal components, genotype batch number, assessment centre, age and sex; Model 2–adjusted for the first six population principal components, genotype batch number, assessment centre, age, sex, highest educational/professional qualification, cohabitation with spouse/partner, annual gross household income, Townsend deprivation index and fasting time. **(A** and **B)** Asterisks indicate statistical significance after correcting *p*-values for multiple testing using the Benjamini–Hochberg procedure. Sample sizes reported in Tables S16 and S17.

### MileAge delta and incident dementia by dementia polygenic scores

About 87% of hazard ratios were below one for individuals with low PRS; 53% of these estimates were statistically significant (Table S18). In the middle PRS group, individuals with a MileAge exceeding chronological age had higher hazards of Alzheimer’s disease, vascular, unspecified and all-cause dementia. Individuals in the high PRS group had higher hazards of dementia (except for dementia in other diseases) across MileAge delta subgroups. The strongest associations were observed for Alzheimer’s disease in those with a high PRS across MileAge delta groups (HRs from 3.69 to 3.94) (Figure 5B).

### Cross-product interactions

We identified only one nominally significant interaction between MileAge delta and *APOE* genotype, suggesting that a one-year higher MileAge delta was associated with 17% higher hazards of dementia in other diseases in the high vs low *APOE* risk groups (Table S19). No statistically significant interactions with dementia PRS were observed (Table S20).

## Discussion

In over 220,000 individuals, a metabolite-predicted age (MileAge) exceeding chronological age was associated with higher hazards of vascular, unspecified and all-cause dementia. Individuals with a MileAge delta greater than one standard deviation above the mean had 20% higher hazards of all-cause dementia than those with a MileAge delta at least one standard deviation below the mean. A higher MileAge delta was also associated with an earlier dementia onset, including of Alzheimer’s disease. Lipids, lipoproteins and amino acids importantly contributed to the MileAge clock and were associated with dementia. Genetic risk factors, including *APOE* genotype and dementia polygenic scores, and MileAge delta were jointly associated with dementia, with little evidence of multiplicative interactions. Individuals with a metabolite-predicted age exceeding their chronological age and a dementia polygenic score greater than one standard deviation above the mean or two copies of the *APOE* ε4 allele had 1.9 to 11-fold higher hazards of vascular dementia.

Our findings broadly align with prior studies linking metabolites (Tynkkynen et al., 2018) and metabolomic profiles (Buergel et al., 2022) to dementia. Our study highlights the role of metabolomic ageing in dementia, especially vascular dementia. The lack of statistically significant association between MileAge delta and Alzheimer’s disease contrasts with studies that identified metabolites linked to Alzheimer’s disease risk (Tynkkynen et al., 2018). Given the role of dysregulated lipid metabolism in Alzheimer’s disease (Yin, 2023), this finding is unexpected. Notably, we identified fewer metabolites associated with Alzheimer’s disease than with other dementia outcomes. PhenoAge, a second-generation ageing clock trained on physiological dysregulation, was associated with Alzheimer’s disease (HR = 1.04 per 1-year increase in PhenoAge) in a prior study of 30 incident cases (Levine et al., 2018). Our metabolome-wide analysis identified branched-chain amino acids as inversely associated with Alzheimer’s disease, which is consistent with prior research (Tynkkynen et al., 2018).

The association between MileAge delta and earlier dementia onset highlights the potential of the MileAge clock to predict not only dementia risk but also timing of disease onset. This is relevant because of the long preclinical period of neurodegenerative diseases. Notably, MileAge delta was associated with an earlier Alzheimer’s disease onset, despite not being associated with incident disease. This could suggest that the MileAge clock captures indicators of speed of progression of Alzheimer’s disease. While this finding warrants cautious interpretation and replication in independent cohorts, it broadly aligns with findings from deCODE genetics showing that proteomic risk scores capture short-term disease processes, while genetic risk scores capture long-term disease risk (Helgason et al., 2023).

Our study also provides insights into the joint association of genetic risk and metabolomic ageing with incident dementia. We did not find that a higher genetic risk (assessed through *APOE* genotype or dementia polygenic scores) was associated with a metabolite-predicted age exceeding chronological age. Instead, both dementia polygenic scores and *APOE* genotype were modestly associated with a MileAge younger than chronological age. Given that genetic risk and MileAge delta were independently associated with dementia, our results indicate that metabolomic ageing does not mediate the relationship between genetic risk and dementia. Rather, genetic risk and MileAge delta appear to represent separate, additive pathways associated with dementia risk.

Our study has certain limitations. Dementia diagnoses were ascertained from routine care and likely include some misclassification. We addressed this through sensitivity analyses excluding self-reported physician diagnosis data and individuals with multiple diagnostic labels—a common occurrence in dementia (Alzheimer’s Association Report, 2023). Some of these concerns could be addressed through changes in diagnostic practice towards biological criteria (Jack Jr. et al., 2024); but see Dubois et al. (2024). To avoid missing dementia cases from primary care, we performed sensitivity analyses in participants with linked primary care records, though with a shorter follow-up. Competing risks were considered in a sensitivity analysis excluding those with multiple diagnostic labels; due to diagnostic uncertainty in dementia (Ranson et al., 2019), no competing risk models were fitted. The coverage of the Nightingale Health platform used to develop the MileAge clock is lipid and lipoprotein focussed, omitting certain metabolites linked to ageing. Most participants only had metabolomics data for a single time point. Longitudinal studies with repeated measures could clarify the temporal relationship between metabolomic ageing and dementia onset. Our study’s observational nature precludes causal inference; reverse causality cannot be ruled out despite our efforts to address it through a sensitivity analysis excluding the first two years of follow-up. While we adjusted for key confounders, residual confounding may persist. Future studies could provide a comparison between MileAge delta and other ageing markers, investigating whether integration with other markers improves dementia risk prediction.

## Conclusion

A metabolite-predicted age that exceeds chronological age was associated with higher hazards of vascular, unspecified and all-cause dementia, as well as earlier dementia onset, including of Alzheimer’s disease. These findings suggest that MileAge delta could help identify individuals at risk before clinical symptoms emerge. Future risk stratification incorporating both genetic and modifiable factors, such as metabolomic age, may enable more targeted preventative strategies.

## Supporting information

Supplementary information is available online.

## Data Availability

The data used are available to all bona fide researchers for health-related research that is in the public interest, subject to an application process and approval criteria. Study materials are publicly available online at http://www.ukbiobank.ac.uk.

## Acknowledgments

JM is funded by the King’s Prize Fellowship and received support through an IoPPN Early Career Research Award. This study is part-funded through the National Institute for Health and Care Research (NIHR) Maudsley Biomedical Research Centre at South London and Maudsley NHS Foundation Trust and King’s College London. The views expressed are those of the authors and not necessarily those of the NHS, the NIHR or the Department of Health and Social Care. LG is funded by the King’s College London DRIVE-Health Centre for Doctoral Training and the Perron Institute for Neurological and Translational Science. OP is supported by a Sir Henry Wellcome Postdoctoral Fellowship [222811/Z/21/Z].

Computational analyses were supported by King’s Computational Research, Engineering and Technology Environment (CREATE). This research has been conducted using data from UK Biobank. Data access permission has been granted under UK Biobank application 45514.

## Authorship contributions

JM conceived the idea of the study, acquired the data, carried out the analysis, interpreted the findings and wrote the manuscript. LG and OP provided methodological support. LG, OP, PP and CML reviewed and edited the manuscript. All authors read and approved the final manuscript.

## Ethics

Ethical approval for the UK Biobank study has been granted by the National Information Governance Board for Health and Social Care and the NHS North West Multicentre Research Ethics Committee (11/NW/0382). No project-specific ethical approval is needed.

## Data sharing statement

The data used are available to all *bona fide* researchers for health-related research that is in the public interest, subject to an application process and approval criteria. Study materials are publicly available online at http://www.ukbiobank.ac.uk.

## Supplementary material

Supplementary information is available online.

