## Supplementary information is available online. for "Accelerated metabolomic ageing (MileAge) in mid-life predicts incident vascular, unspecified and all-cause dementia"

This material accompanies the article

### 1. Supplementary methods

**Imputed genotype data**

Details on the genotype calling, quality control and imputation performed centrally by the UK Biobank have been reported elsewhere (Bycroft et al., 2018). Individuals were genotyped using the UK BiLEVE Axiom ($N$ = 49,950) and UK Biobank Axiom ($N$ = 438,427) arrays. The data were imputed using the Haplotype Reference Consortium (HRC) and the combined UK10K and 1000 Genomes Project phase 3 reference panels, with imputed data available for approximately 93,095,623 autosomal SNPs. These were filtered using a minor allele frequency (MAF) threshold of $\geq$ 1% and INFO score of $\geq$ 0.4.

**Genotype quality control**

Genotype quality control steps were applied using the GenoPred pipeline (v2.2.10) (Pain, Al-Chalabi, & Lewis, 2024).

<u>GWAS summary statistics</u>: Variants were excluded if they were not identified in the reference dataset, were strand ambiguous, had an INFO score < 0.9 (if available), had out-of-bound $p$-values ($0 < P \leq 1$), had duplicate SNP IDs, had a sample size >3SD from the median (if available) or had a standard error of zero. MAF thresholds were set at 0.01 for the GWAS summary statistics and the reference dataset, with a MAF difference threshold of 0.2.

<u>Individual-level genotype data</u>: Variants absent from the reference data and duplicate variants were removed. Variants with mismatched RSIDs and variants that were strand-flipped were corrected. Strand-ambiguous variants were excluded. Reference variants absent from the UK Biobank data were inserted as missing to allow reference allele frequency-based imputation during downstream polygenic scoring. Relatedness was estimated using the KING estimator in PLINK v1.9, with unrelated individuals defined using a threshold of $r$ > 0.044 (equivalent to removing third-degree relatives and closer).

#### 2. UK Biobank data fields

**Table S1**. UK Biobank data

| Data field IDs | Variable name |
|---|---|
| 31 | Sex |
| 34 | Year of birth |
| 52 | Month of birth |
| 53 | Date of attending assessment centre |
| 54 | UK Biobank assessment centre |
| 74 | Fasting time |
| 137 | Number of treatments/medications taken |
| 191 | Date lost to follow-up |
| 738 | Average total household income before tax |
| 3140 | Pregnant |
| 6138[1] | Qualifications |
| 6141[1] | How are people in household related to participant |
| 6153 | Medication for cholesterol, blood pressure, diabetes, or take exogenous hormones |
| 6177 | Medication for cholesterol, blood pressure or diabetes |
| 20003 | Treatment/medication code |
| 21000[1] | Ethnic background |
| 22000 | Genotype measurement batch |
| 22001 | Genetic sex |
| 22009 | Genetic principal components |
| 22189[1] | Townsend deprivation index at recruitment |
| 40000 | Date of death |
| 130836 | Date F00 first reported (dementia in alzheimer's disease) |
| 130837 | Source of report of F00 (dementia in alzheimer's disease) |
| 130838 | Date F01 first reported (vascular dementia) |
| 130839 | Source of report of F01 (vascular dementia) |
| 130840 | Date F02 first reported (dementia in other diseases classified elsewhere) |
| 130841 | Source of report of F02 (dementia in other diseases classified elsewhere) |
| 130842 | Date F03 first reported (unspecified dementia) |
| 130843 | Source of report of F03 (unspecified dementia) |
| 131036 | Date G30 first reported (alzheimer's disease) |
| 131037 | Source of report of G30 (alzheimer's disease) |

*Note:* [1]These variables were further processed and categories used in the present study are shown in the main body of the text.

Details on the data fields for the Nightingale Health nuclear magnetic resonance (NMR) spectroscopy metabolomic biomarkers can be found in the UK Biobank showcase:

https://biobank.ndph.ox.ac.uk/showcase/label.cgi?id=220

#### 3. GWAS summary statistics

**Table S2.** Genome-wide association study (GWAS) summary statistics

| | $N_{\text{Cases}}$ | $N_{\text{Controls}}$ | $N_{\text{Total}}$ | $N_{\text{Eff.}}$ | Sampling | Prevalence | Citation | PMID | UKB |
|---|---|---|---|---|---|---|---|---|---|
| Alzheimer's disease | 39918 | 358140 | 398058 | 115084 | 0.10028190 | 0.070 | Wightman et al., 2021* | 34493870; 37045620 | No |
| Vascular dementia | 3624 | 475484 | 479108 | 14386 | 0.00756406 | 0.015 | Kurki et al., 2023 | 36653562 | No |
| Dementia in other diseases | 1607 | 469981 | 471588 | 6407 | 0.00340764 | 0.003 | Kurki et al., 2023 | 36653562 | No |
| Dementia (unspecified) | 5388 | 469981 | 475369 | 21306 | 0.01133435 | 0.020 | Kurki et al., 2023 | 36653562 | No |
| All-cause dementia | 24864 | 469981 | 494845 | 94460 | 0.05024604 | 0.064 | Kurki et al., 2023 | 36653562 | No |

*Note:* PMID = PubMed reference number; UKB = UK Biobank. * excluding UKB and 23andMe.

4. Directed acyclic graph

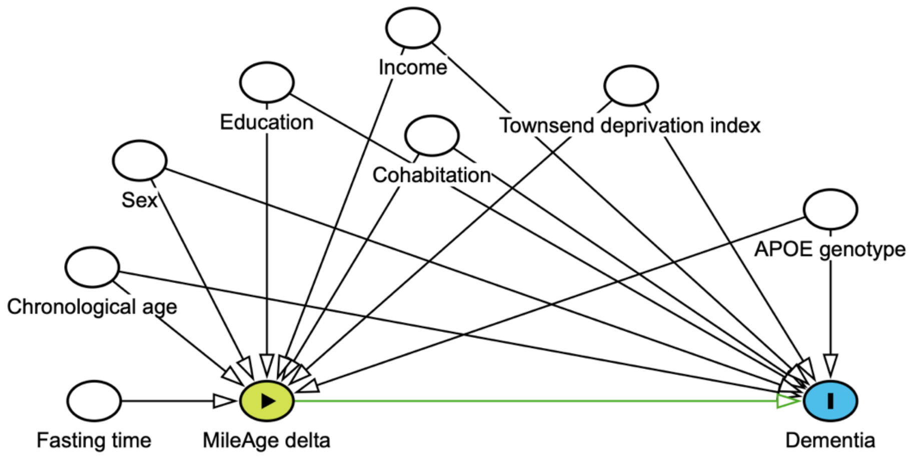

**Figure S1.** Directed acyclic graph (DAG) showing relationships between the exposure (MileAge delta), outcome (incident dementia) and select confounders (age, sex, highest educational/professional qualification, cohabitation with spouse/partner, annual gross household income, Townsend deprivation index and fasting time).

#### 5. Incident dementia by ethnicity

**Table S3.** Incident dementia by ethnicity

| | *N* total | *N* incident |
|---|---|---|
| Alzheimer's disease | | |
| White | 209797 | 1806 |
| Mixed | 1238 | 8 |
| Black | 3130 | 27 |
| Asian | 3796 | 23 |
| Chinese | 631 | 0 |
| Other | 1834 | 6 |
| Missing[1] | 975 | 11 |
| Vascular dementia | | |
| White | 208901 | 910 |
| Mixed | 1233 | 3 |
| Black | 3111 | 8 |
| Asian | 3780 | 7 |
| Chinese | 631 | 0 |
| Other | 1832 | 4 |
| Missing[1] | 965 | 1 |
| Dementia in other diseases | | |
| White | 208482 | 491 |
| Mixed | 1231 | 1 |
| Black | 3112 | 9 |
| Asian | 3778 | 5 |
| Chinese | 632 | 1 |
| Other | 1831 | 3 |
| Missing[1] | 966 | 2 |
| Dementia (unspecified) | | |
| White | 210141 | 2150 |
| Mixed | 1238 | 8 |
| Black | 3130 | 27 |
| Asian | 3799 | 26 |
| Chinese | 633 | 2 |
| Other | 1838 | 10 |
| Missing[1] | 981 | 17 |
| All-cause dementia | | |
| White | 211822 | 3831 |
| Mixed | 1245 | 15 |
| Black | 3148 | 45 |
| Asian | 3816 | 43 |
| Chinese | 633 | 2 |
| Other | 1844 | 16 |
| Missing[1] | 988 | 24 |

*Note:* [1] Missing data may also include "do not know" or "prefer not to answer".

#### 6. Lipid-modifying treatment codes

**Table S4.** Lipid-modifying treatment codes

| Code | Meaning |
|---|---|
| 1193 | Omega-3/fish oil supplement |
| 1140861848 | Simvastatin 10mg tablet |
| 1140861856 | Simvastatin 20mg tablet |
| 1140861892 | Simvastatin 40mg tablet |
| 1140861922 | Atorvastatin 10mg tablet |
| 1140861924 | Atorvastatin 20mg tablet |
| 1140861926 | Atorvastatin 40mg tablet |
| 1140861928 | Atorvastatin 80mg tablet |
| 1140861936 | Pravastatin 10mg tablet |
| 1140861944 | Pravastatin 20mg tablet |
| 1140861954 | Pravastatin 40mg tablet |
| 1140861958 | Fluvastatin 20mg capsule |
| 1140861970 | Fluvastatin 40mg capsule |
| 1140862026 | Rosuvastatin 5mg tablet |
| 1140862028 | Rosuvastatin 10mg tablet |
| 1140864592 | Ezetimibe 10mg tablet |
| 1140865576 | Simvastatin 80mg tablet |
| 1140881748 | Bezafibrate 200mg tablet |
| 1140888590 | Fenofibrate 200mg capsule |
| 1140888594 | Fenofibrate 267mg capsule |
| 1140888648 | Gemfibrozil 600mg tablet |
| 1140909780 | Colestyramine 4g/sachet powder |
| 1141146138 | Atorvastatin 30mg tablet |
| 1141146234 | Rosuvastatin 20mg tablet |
| 1141157260 | Ezetimibe/simvastatin 10mg/20mg tablet |
| 1141162544 | Ezetimibe/simvastatin 10mg/40mg tablet |
| 1141171548 | Ezetimibe/atorvastatin 10mg/10mg tablet |
| 1141172214 | Ezetimibe/atorvastatin 10mg/20mg tablet |
| 1141180722 | Ezetimibe/atorvastatin 10mg/40mg tablet |
| 1141180734 | Ezetimibe/atorvastatin 10mg/80mg tablet |
| 1141181868 | Pitavastatin 1mg tablet |
| 1141188146 | Pitavastatin 2mg tablet |
| 1141188546 | Pitavastatin 4mg tablet |
| 1141192410 | Alirocumab 75mg/ml solution for injection pre-filled pen |
| 1141192414 | Alirocumab 150mg/ml solution for injection pre-filled pen |
| 1141192736 | Evolocumab 140mg/ml solution for injection pre-filled pen |
| 1141192740 | Evolocumab 140mg/ml solution for injection pre-filled syringe |
| 1141195196 | Bempedoic acid 180mg tablet |
| 1141200040 | Bempedoic acid/ezetimibe 180mg/10mg tablet |
| 1141201306 | Inclisiran 284mg/1.5ml solution for injection pre-filled syringe |

*Note:* Participants reported any regular treatments taken, including prescription medications, over-the-counter medications, vitamins and supplements.

#### 7. Incident cases by data source

**Table S5.** Incident cases by data source

| | Death | | Primary care | | Hospital | | Self-report | |
|---|---|---|---|---|---|---|---|---|
| | Only | + | Only | + | Only | + | Only | + |
| Alzheimer's disease | 119 | 0 | 18 | 15 | 1684 | 45 | 0 | 0 |
| Vascular dementia | 54 | 1 | 41 | 50 | 620 | 167 | 0 | 0 |
| Dementia in other diseases | 0 | 0 | 5 | 1 | 506 | 0 | 0 | 0 |
| Dementia (unspecified) | 261 | 0 | 12 | 23 | 1595 | 333 | 12 | 4 |

*Note:* Incident cases were ascertained from coded primary care data, hospital inpatient data, death registry data and participant self-report data of medical diagnoses. Details can be found under category 1712 in the UK Biobank showcase.

8. MileAge delta and incident dementia, excluding self-report data

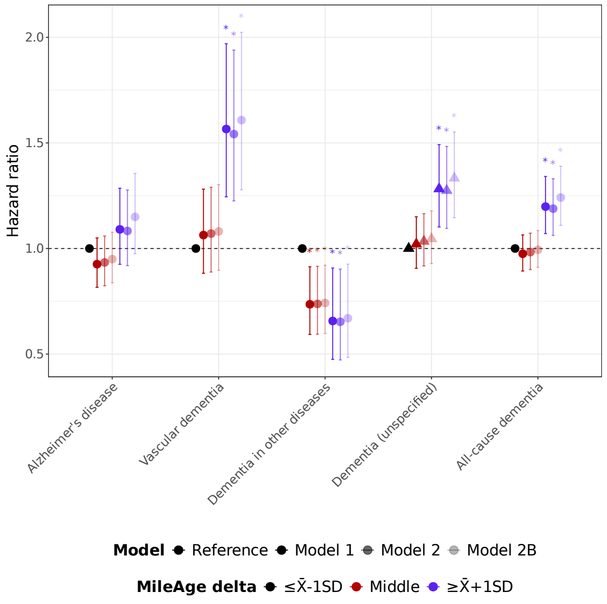

**Figure S2**. Hazard ratios (HR) and 95% confidence intervals from Cox proportional hazards models for incident dementia by MileAge delta, excluding self-report data of medical diagnoses. Time since the baseline assessment (in days) was used as the underlying time axis. Reference group: individuals with a MileAge delta smaller than one standard deviation below the mean. Model 1–adjusted for age and sex; Model 2–adjusted for age, sex, highest educational/professional qualification, cohabitation with spouse/partner, annual gross household income, Townsend deprivation index and fasting time; Model 2B–additionally adjusted for apolipoprotein E (*APOE*) genotype. Asterisks indicate statistical significance after correcting *p*-values for multiple testing using the Benjamini–Hochberg procedure. *N* = 1881 (Alzheimer's disease); *N* = 933 (vascular dementia); *N* = 512 (dementia in other diseases); *N* = 2224 (unspecified dementia); *N* = 3976 (all-cause dementia).

**Table S6.** MileAge delta and incident dementia, excluding self-report data

| | | | Model 1 | | | | Model 2 | | | | Model 2B | | | |
|---|---|---|---|---|---|---|---|---|---|---|---|---|---|---|
| Level | $N_{\text{total}}$ | $N_{\text{incident}}$ | HR | 95% CI | | *p* | HR | 95% CI | | *p* | HR | 95% CI | | *p* |
| Dementia (unspecified) | | | | | | | | | | | | | | |
| ≤ X̄-1SD | 35686 | 331 | Reference | | | | | | | | | | | |
| Middle | 150987 | 1548 | 1.02 | 0.91 | 1.15 | 0.758 | 1.03 | 0.92 | 1.16 | 0.650 | 1.05 | 0.93 | 1.18 | 0.591 |
| ≥ X̄+1SD | 35071 | 345 | 1.28 | 1.10 | 1.49 | 0.006 | 1.27 | 1.10 | 1.48 | 0.006 | 1.33 | 1.15 | 1.55 | 0.001 |

*Note:* HR = hazard ratio; CI = confidence interval; SD = standard deviation. Time since the baseline assessment (in days) was used as the underlying time axis. Model 1–adjusted for age and sex; Model 2–adjusted for age, sex, highest educational/professional qualification, cohabitation with spouse/partner, annual gross household income, Townsend deprivation index and fasting time; Model 2B–additionally adjusted for apolipoprotein E (*APOE*) genotype. *P*-values shown are corrected for multiple testing using the Benjamini–Hochberg procedure (across all outcomes, exposure levels and models).

#### 9. MileAge delta and incident dementia, excluding the first two years

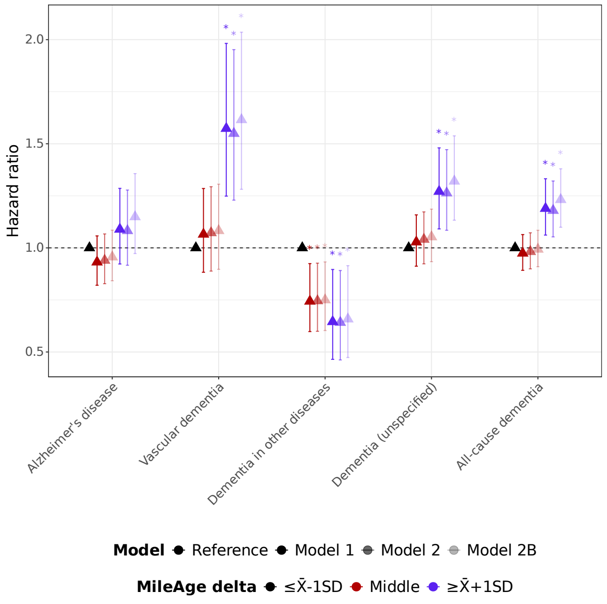

**Figure S3**. Hazard ratios (HR) and 95% confidence intervals from Cox proportional hazards models for incident dementia by MileAge delta, excluding the first two years of follow-up. Time since the baseline assessment (in days) was used as the underlying time axis. Reference group: individuals with a MileAge delta smaller than one standard deviation below the mean. Model 1–adjusted for age and sex; Model 2–adjusted for age, sex, highest educational/professional qualification, cohabitation with spouse/partner, annual gross household income, Townsend deprivation index and fasting time; Model 2B–additionally adjusted for apolipoprotein E (*APOE*) genotype. Asterisks indicate statistical significance after correcting *p*-values for multiple testing using the Benjamini–Hochberg procedure. $N$ = 1858 (Alzheimer's disease); $N$ = 920 (vascular dementia); $N$ = 505 (dementia in other diseases); $N$ = 2211 (unspecified dementia); $N$ = 3930 (all-cause dementia).

**Table S7.** MileAge delta and incident dementia, excluding first two years

| | | | Model 1 | | | | Model 2 | | | | Model 2B | | | |
|---|---|---|---|---|---|---|---|---|---|---|---|---|---|---|
| Level | $N_{total}$ | $N_{incident}$ | HR | 95% CI | | *p* | HR | 95% CI | | *p* | HR | 95% CI | | *p* |
| **Alzheimer's disease** | | | | | | | | | | | | | | |
| ≤ X̄-1SD | 35526 | 296 | Reference | | | | | | | | | | | |
| Middle | 150052 | 1293 | 0.93 | 0.82 | 1.06 | 0.483 | 0.94 | 0.83 | 1.07 | 0.525 | 0.96 | 0.84 | 1.09 | 0.592 |
| ≥ X̄+1SD | 34757 | 269 | 1.09 | 0.92 | 1.29 | 0.521 | 1.08 | 0.92 | 1.28 | 0.525 | 1.15 | 0.97 | 1.36 | 0.191 |
| **Vascular dementia** | | | | | | | | | | | | | | |
| ≤ X̄-1SD | 35363 | 133 | Reference | | | | | | | | | | | |
| Middle | 149386 | 627 | 1.07 | 0.88 | 1.29 | 0.592 | 1.07 | 0.89 | 1.29 | 0.592 | 1.08 | 0.90 | 1.31 | 0.556 |
| ≥ X̄+1SD | 34648 | 160 | 1.57 | 1.25 | 1.98 | 0.002 | 1.55 | 1.23 | 1.95 | 0.002 | 1.62 | 1.28 | 2.04 | 0.001 |
| **Dementia in other diseases** | | | | | | | | | | | | | | |
| ≤ X̄-1SD | 35337 | 107 | Reference | | | | | | | | | | | |
| Middle | 149103 | 344 | 0.74 | 0.60 | 0.92 | 0.020 | 0.75 | 0.60 | 0.93 | 0.020 | 0.75 | 0.60 | 0.93 | 0.021 |
| ≥ X̄+1SD | 34542 | 54 | 0.65 | 0.46 | 0.90 | 0.021 | 0.64 | 0.46 | 0.89 | 0.020 | 0.66 | 0.47 | 0.91 | 0.025 |
| **Dementia (unspecified)** | | | | | | | | | | | | | | |
| ≤ X̄-1SD | 35558 | 328 | Reference | | | | | | | | | | | |
| Middle | 150303 | 1544 | 1.03 | 0.91 | 1.16 | 0.697 | 1.04 | 0.92 | 1.17 | 0.592 | 1.05 | 0.93 | 1.19 | 0.556 |
| ≥ X̄+1SD | 34827 | 339 | 1.27 | 1.09 | 1.48 | 0.010 | 1.26 | 1.08 | 1.47 | 0.010 | 1.32 | 1.13 | 1.54 | 0.002 |
| **All-cause dementia** | | | | | | | | | | | | | | |
| ≤ X̄-1SD | 35842 | 612 | Reference | | | | | | | | | | | |
| Middle | 151483 | 2724 | 0.97 | 0.89 | 1.06 | 0.624 | 0.98 | 0.90 | 1.07 | 0.710 | 0.99 | 0.91 | 1.08 | 0.880 |
| ≥ X̄+1SD | 35082 | 594 | 1.19 | 1.06 | 1.33 | 0.010 | 1.18 | 1.05 | 1.32 | 0.015 | 1.23 | 1.10 | 1.38 | 0.002 |

*Note:* HR = hazard ratio; CI = confidence interval; SD = standard deviation. Time since the baseline assessment (in days) was used as the underlying time axis. Model 1–adjusted for age and sex; Model 2–adjusted for age, sex, highest educational/professional qualification, cohabitation with spouse/partner, annual gross household income, Townsend deprivation index and fasting time; Model 2B–additionally adjusted for apolipoprotein E (*APOE*) genotype. *P*-values shown are corrected for multiple testing using the Benjamini–Hochberg procedure (across all outcomes, exposure levels and models).

10. MileAge delta and incident dementia, individuals aged ≥ 60 years

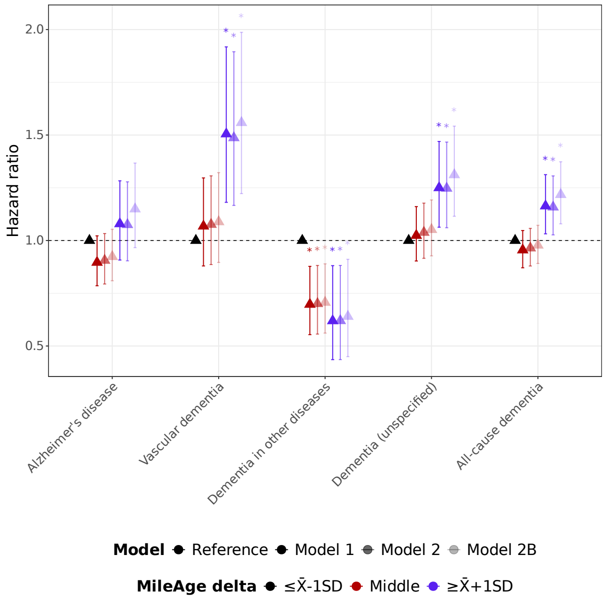

**Figure S4**. Hazard ratios (HR) and 95% confidence intervals from Cox proportional hazards models for incident dementia by MileAge delta, in individuals aged ≥ 60 years at the baseline assessment. Time since the baseline assessment (in days) was used as the underlying time axis. Reference group: individuals with a MileAge delta smaller than one standard deviation below the mean. Model 1–adjusted for age and sex; Model 2–adjusted for age, sex, highest educational/professional qualification, cohabitation with spouse/partner, annual gross household income, Townsend deprivation index and fasting time; Model 2B–additionally adjusted for apolipoprotein E (*APOE*) genotype. Asterisks indicate statistical significance after correcting *p*-values for multiple testing using the Benjamini–Hochberg procedure. *N* = 1699 (Alzheimer's disease); *N* = 859 (vascular dementia); *N* = 441 (dementia in other diseases); *N* = 2006 (unspecified dementia); *N* = 3548 (all-cause dementia).

**Table S8.** MileAge delta and incident dementia, individuals aged ≥ 60 years

| Level | *N* total | *N* incident | Model 1 HR | Model 1 95% CI | | Model 1 *p* | Model 2 HR | Model 2 95% CI | | Model 2 *p* | Model 2B HR | Model 2B 95% CI | | Model 2B *p* |
|---|---|---|---|---|---|---|---|---|---|---|---|---|---|---|
| Alzheimer's disease | | | | | | | | | | | | | | |
| ≤ X̄-1SD | 13912 | 276 | Reference | | | | | | | | | | | |
| Middle | 67343 | 1179 | 0.90 | 0.79 | 1.02 | 0.192 | 0.91 | 0.79 | 1.03 | 0.233 | 0.92 | 0.81 | 1.05 | 0.371 |
| ≥ X̄+1SD | 13139 | 244 | 1.08 | 0.91 | 1.28 | 0.528 | 1.07 | 0.90 | 1.28 | 0.528 | 1.15 | 0.97 | 1.37 | 0.204 |
| Vascular dementia | | | | | | | | | | | | | | |
| ≤ X̄-1SD | 13760 | 124 | Reference | | | | | | | | | | | |
| Middle | 66758 | 594 | 1.07 | 0.88 | 1.30 | 0.563 | 1.08 | 0.89 | 1.31 | 0.531 | 1.09 | 0.90 | 1.32 | 0.528 |
| ≥ X̄+1SD | 13036 | 141 | 1.51 | 1.18 | 1.92 | 0.008 | 1.49 | 1.17 | 1.89 | 0.008 | 1.56 | 1.22 | 1.99 | 0.008 |
| Dementia in other diseases | | | | | | | | | | | | | | |
| ≤ X̄-1SD | 13733 | 97 | Reference | | | | | | | | | | | |
| Middle | 66462 | 298 | 0.70 | 0.55 | 0.88 | 0.010 | 0.70 | 0.56 | 0.88 | 0.010 | 0.71 | 0.56 | 0.89 | 0.012 |
| ≥ X̄+1SD | 12941 | 46 | 0.62 | 0.44 | 0.88 | 0.020 | 0.62 | 0.44 | 0.88 | 0.020 | 0.64 | 0.45 | 0.91 | 0.029 |
| Dementia (unspecified) | | | | | | | | | | | | | | |
| ≤ X̄-1SD | 13933 | 297 | Reference | | | | | | | | | | | |
| Middle | 67578 | 1414 | 1.02 | 0.90 | 1.16 | 0.714 | 1.04 | 0.92 | 1.18 | 0.598 | 1.05 | 0.93 | 1.19 | 0.528 |
| ≥ X̄+1SD | 13190 | 295 | 1.25 | 1.06 | 1.47 | 0.020 | 1.25 | 1.06 | 1.47 | 0.020 | 1.31 | 1.11 | 1.54 | 0.008 |
| All-cause dementia | | | | | | | | | | | | | | |
| ≤ X̄-1SD | 14193 | 557 | Reference | | | | | | | | | | | |
| Middle | 68637 | 2473 | 0.95 | 0.87 | 1.05 | 0.492 | 0.96 | 0.88 | 1.06 | 0.528 | 0.98 | 0.89 | 1.07 | 0.652 |
| ≥ X̄+1SD | 13413 | 518 | 1.16 | 1.03 | 1.31 | 0.029 | 1.16 | 1.03 | 1.31 | 0.034 | 1.22 | 1.08 | 1.37 | 0.008 |

*Note:* HR = hazard ratio; CI = confidence interval; SD = standard deviation. Time since the baseline assessment (in days) was used as the underlying time axis. Model 1–adjusted for age and sex; Model 2–adjusted for age, sex, highest educational/professional qualification, cohabitation with spouse/partner, annual gross household income, Townsend deprivation index and fasting time; Model 2B–additionally adjusted for apolipoprotein E (*APOE*) genotype. *P*-values shown are corrected for multiple testing using the Benjamini–Hochberg procedure (across all outcomes, exposure levels and models).

#### 11. Sample flowchart, primary care linkage

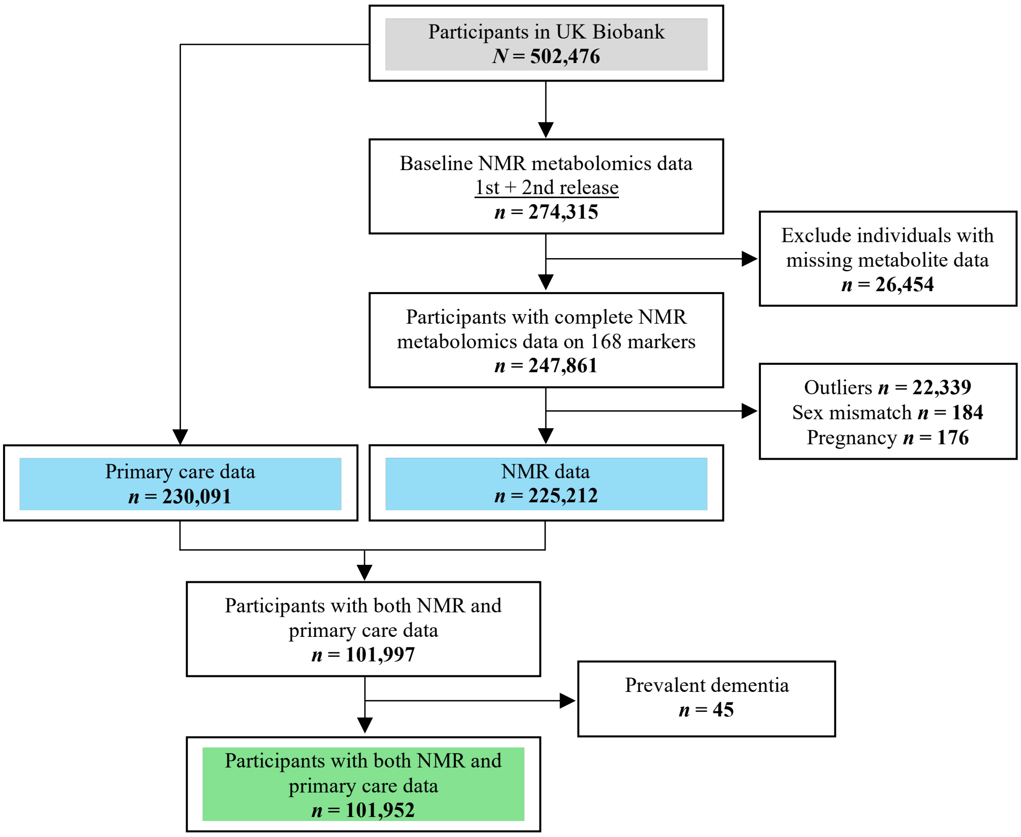

**Figure S5**. Study sample flowchart. Outliers were defined as metabolite values 4×IQR above or below the median. NMR = nuclear magnetic resonance; IQR = interquartile range.

#### 12. MileAge delta and incident dementia, primary care linkage

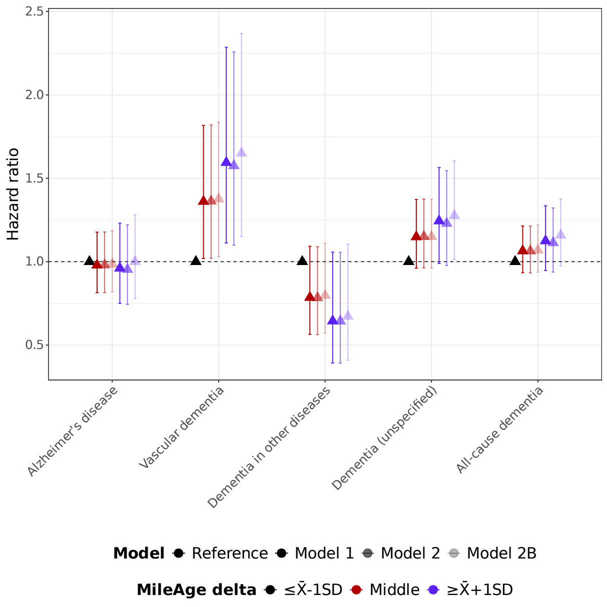

**Figure S6**. Hazard ratios (HR) and 95% confidence intervals from Cox proportional hazards models for incident dementia by MileAge delta, in individuals with primary care linkage. Time since the baseline assessment (in days) was used as the underlying time axis. Reference group: individuals with a MileAge delta smaller than one standard deviation below the mean. Model 1–adjusted for age and sex; Model 2–adjusted for age, sex, highest educational/professional qualification, cohabitation with spouse/partner, annual gross household income, Townsend deprivation index and fasting time; Model 2B–additionally adjusted for apolipoprotein E (*APOE*) genotype. Asterisks indicate statistical significance after correcting *p*-values for multiple testing using the Benjamini–Hochberg procedure. *N* = 888 (Alzheimer's disease); *N* = 442 (vascular dementia); *N* = 226 (dementia in other diseases); *N* = 1054 (unspecified dementia); *N* = 1836 (all-cause dementia).

**Table S9.** MileAge delta and incident dementia, primary care linkage

| Level | $N_{total}$ | $N_{incident}$ | Model 1 HR | Model 1 95% CI | | Model 1 *p* | Model 2 HR | Model 2 95% CI | | Model 2 *p* | Model 2B HR | Model 2B 95% CI | | Model 2B *p* |
|---|---|---|---|---|---|---|---|---|---|---|---|---|---|---|
| Alzheimer's disease | | | | | | | | | | | | | | |
| ≤ X̄-1SD | 15895 | 139 | Reference | | | | | | | | | | | |
| Middle | 68799 | 633 | 0.98 | 0.81 | 1.18 | 0.887 | 0.98 | 0.81 | 1.18 | 0.887 | 0.99 | 0.82 | 1.19 | 0.907 |
| ≥ X̄+1SD | 16310 | 116 | 0.96 | 0.75 | 1.23 | 0.866 | 0.95 | 0.74 | 1.22 | 0.844 | 1.00 | 0.78 | 1.28 | >0.999 |
| Vascular dementia | | | | | | | | | | | | | | |
| ≤ X̄-1SD | 15810 | 54 | Reference | | | | | | | | | | | |
| Middle | 68487 | 321 | 1.36 | 1.02 | 1.82 | 0.161 | 1.36 | 1.02 | 1.82 | 0.161 | 1.38 | 1.03 | 1.84 | 0.161 |
| ≥ X̄+1SD | 16261 | 67 | 1.59 | 1.11 | 2.29 | 0.135 | 1.57 | 1.10 | 2.26 | 0.135 | 1.65 | 1.15 | 2.37 | 0.135 |
| Dementia in other diseases | | | | | | | | | | | | | | |
| ≤ X̄-1SD | 15802 | 46 | Reference | | | | | | | | | | | |
| Middle | 68322 | 156 | 0.79 | 0.56 | 1.09 | 0.252 | 0.78 | 0.56 | 1.09 | 0.252 | 0.80 | 0.57 | 1.11 | 0.271 |
| ≥ X̄+1SD | 16218 | 24 | 0.64 | 0.39 | 1.06 | 0.223 | 0.64 | 0.39 | 1.06 | 0.223 | 0.67 | 0.41 | 1.10 | 0.240 |
| Dementia (unspecified) | | | | | | | | | | | | | | |
| ≤ X̄-1SD | 15901 | 145 | Reference | | | | | | | | | | | |
| Middle | 68924 | 758 | 1.15 | 0.96 | 1.37 | 0.240 | 1.15 | 0.96 | 1.38 | 0.240 | 1.15 | 0.96 | 1.37 | 0.240 |
| ≥ X̄+1SD | 16345 | 151 | 1.24 | 0.99 | 1.56 | 0.223 | 1.23 | 0.98 | 1.55 | 0.223 | 1.28 | 1.01 | 1.60 | 0.161 |
| All-cause dementia | | | | | | | | | | | | | | |
| ≤ X̄-1SD | 16026 | 270 | Reference | | | | | | | | | | | |
| Middle | 69476 | 1310 | 1.06 | 0.93 | 1.21 | 0.446 | 1.06 | 0.93 | 1.21 | 0.446 | 1.07 | 0.94 | 1.22 | 0.437 |
| ≥ X̄+1SD | 16450 | 256 | 1.12 | 0.95 | 1.34 | 0.271 | 1.11 | 0.94 | 1.32 | 0.316 | 1.16 | 0.97 | 1.38 | 0.237 |

*Note:* HR = hazard ratio; CI = confidence interval; SD = standard deviation. Time since the baseline assessment (in days) was used as the underlying time axis. Model 1–adjusted for age and sex; Model 2–adjusted for age, sex, highest educational/professional qualification, cohabitation with spouse/partner, annual gross household income, Townsend deprivation index and fasting time; Model 2B–additionally adjusted for apolipoprotein E (*APOE*) genotype. *P*-values shown are corrected for multiple testing using the Benjamini–Hochberg procedure (across all outcomes, exposure levels and models).

### 13. MileAge delta and incident dementia, excluding multiple diagnoses

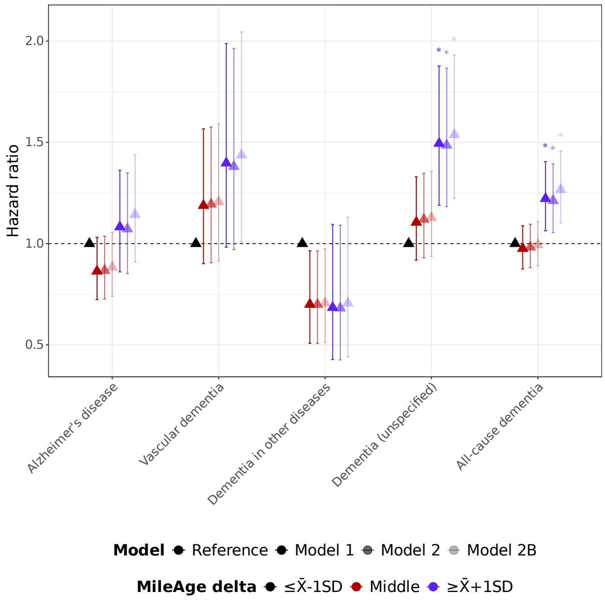

**Figure S7**. Hazard ratios (HR) and 95% confidence intervals from Cox proportional hazards models for incident dementia by MileAge delta, excluding multiple diagnoses. Time since the baseline assessment (in days) was used as the underlying time axis. Reference group: individuals with a MileAge delta smaller than one standard deviation below the mean. Model 1–adjusted for age and sex; Model 2–adjusted for age, sex, highest educational/professional qualification, cohabitation with spouse/partner, annual gross household income, Townsend deprivation index and fasting time; Model 2B–additionally adjusted for apolipoprotein E (*APOE*) genotype. Asterisks indicate statistical significance after correcting *p*-values for multiple testing using the Benjamini–Hochberg procedure. *N* = 920 (Alzheimer's disease); *N* = 441 (vascular dementia); *N* = 229 (dementia in other diseases); *N* = 988 (unspecified dementia); *N* = 2578 (all-cause dementia).

**Table S10.** MileAge delta and incident dementia, excluding multiple diagnoses

| | | | Model 1 | | | | Model 2 | | | | Model 2B | | | |
|---|---|---|---|---|---|---|---|---|---|---|---|---|---|---|
| Level | $N_{total}$ | $N_{incident}$ | HR | 95% CI | | *p* | HR | 95% CI | | *p* | HR | 95% CI | | *p* |
| Alzheimer's disease | | | | | | | | | | | | | | |
| ≤ X̄-1SD | 35508 | 153 | Reference | | | | | | | | | | | |
| Middle | 150064 | 625 | 0.86 | 0.72 | 1.03 | 0.220 | 0.87 | 0.73 | 1.04 | 0.220 | 0.88 | 0.74 | 1.06 | 0.288 |
| ≥ X̄+1SD | 34868 | 142 | 1.08 | 0.86 | 1.36 | 0.575 | 1.07 | 0.85 | 1.35 | 0.612 | 1.14 | 0.91 | 1.44 | 0.317 |
| Vascular dementia | | | | | | | | | | | | | | |
| ≤ X̄-1SD | 35415 | 60 | Reference | | | | | | | | | | | |
| Middle | 149755 | 316 | 1.19 | 0.90 | 1.57 | 0.302 | 1.19 | 0.91 | 1.58 | 0.297 | 1.21 | 0.91 | 1.59 | 0.291 |
| ≥ X̄+1SD | 34791 | 65 | 1.40 | 0.98 | 1.99 | 0.170 | 1.38 | 0.97 | 1.96 | 0.183 | 1.44 | 1.01 | 2.05 | 0.132 |
| Dementia in other diseases | | | | | | | | | | | | | | |
| ≤ X̄-1SD | 35405 | 50 | Reference | | | | | | | | | | | |
| Middle | 149591 | 152 | 0.70 | 0.51 | 0.96 | 0.110 | 0.70 | 0.51 | 0.96 | 0.110 | 0.71 | 0.51 | 0.97 | 0.113 |
| ≥ X̄+1SD | 34753 | 27 | 0.68 | 0.43 | 1.09 | 0.220 | 0.68 | 0.43 | 1.09 | 0.220 | 0.71 | 0.44 | 1.13 | 0.260 |
| Dementia (unspecified) | | | | | | | | | | | | | | |
| ≤ X̄-1SD | 35490 | 135 | Reference | | | | | | | | | | | |
| Middle | 150125 | 686 | 1.11 | 0.92 | 1.33 | 0.347 | 1.12 | 0.93 | 1.35 | 0.302 | 1.13 | 0.94 | 1.36 | 0.297 |
| ≥ X̄+1SD | 34893 | 167 | 1.49 | 1.19 | 1.88 | 0.007 | 1.49 | 1.18 | 1.87 | 0.007 | 1.54 | 1.22 | 1.93 | 0.007 |
| All-cause dementia | | | | | | | | | | | | | | |
| ≤ X̄-1SD | 35753 | 398 | Reference | | | | | | | | | | | |
| Middle | 151218 | 1779 | 0.98 | 0.88 | 1.09 | 0.707 | 0.98 | 0.88 | 1.10 | 0.778 | 0.99 | 0.89 | 1.11 | 0.914 |
| ≥ X̄+1SD | 35127 | 401 | 1.22 | 1.06 | 1.41 | 0.028 | 1.21 | 1.05 | 1.39 | 0.034 | 1.27 | 1.10 | 1.46 | 0.007 |

*Note:* HR = hazard ratio; CI = confidence interval; SD = standard deviation. Time since the baseline assessment (in days) was used as the underlying time axis. Model 1–adjusted for age and sex; Model 2–adjusted for age, sex, highest educational/professional qualification, cohabitation with spouse/partner, annual gross household income, Townsend deprivation index and fasting time; Model 2B–additionally adjusted for apolipoprotein E (*APOE*) genotype. *P*-values shown are corrected for multiple testing using the Benjamini–Hochberg procedure (across all outcomes, exposure levels and models).

#### 14. MileAge delta and incident dementia, excluding lipid-modifying treatments

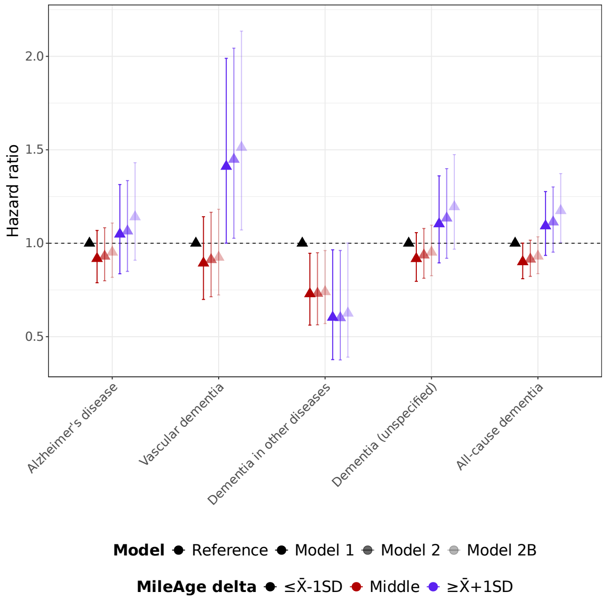

**Figure S8**. Hazard ratios (HR) and 95% confidence intervals from Cox proportional hazards models for incident dementia by MileAge delta, excluding individuals using lipid-modifying treatments. Time since the baseline assessment (in days) was used as the underlying time axis. Reference group: individuals with a MileAge delta smaller than one standard deviation below the mean. Model 1–adjusted for age and sex; Model 2–adjusted for age, sex, highest educational/professional qualification, cohabitation with spouse/partner, annual gross household income, Townsend deprivation index and fasting time; Model 2B–additionally adjusted for apolipoprotein E (*APOE*) genotype. Asterisks indicate statistical significance after correcting *p*-values for multiple testing using the Benjamini–Hochberg procedure. *N* = 1131 (Alzheimer's disease); *N* = 424 (vascular dementia); *N* = 311 (dementia in other diseases); *N* = 1281 (unspecified dementia); *N* = 2283 (all-cause dementia).

**Table S11.** MileAge delta and incident dementia, excluding lipid-modifying treatments

| | | | Model 1 | | | | Model 2 | | | | Model 2B | | | |
|---|---|---|---|---|---|---|---|---|---|---|---|---|---|---|
| Level | $N_{\text{total}}$ | $N_{\text{incident}}$ | HR | 95% CI | | *p* | HR | 95% CI | | *p* | HR | 95% CI | | *p* |
| Alzheimer's disease | | | | | | | | | | | | | | |
| ≤ X̄-1SD | 31702 | 215 | References | | | | | | | | | | | |
| Middle | 121230 | 795 | 0.92 | 0.79 | 1.07 | 0.403 | 0.93 | 0.80 | 1.08 | 0.461 | 0.95 | 0.82 | 1.11 | 0.569 |
| ≥ X̄+1SD | 24333 | 121 | 1.05 | 0.84 | 1.31 | 0.681 | 1.07 | 0.85 | 1.34 | 0.605 | 1.14 | 0.91 | 1.43 | 0.403 |
| Vascular dementia | | | | | | | | | | | | | | |
| ≤ X̄-1SD | 31571 | 84 | References | | | | | | | | | | | |
| Middle | 120719 | 284 | 0.89 | 0.70 | 1.14 | 0.461 | 0.91 | 0.71 | 1.17 | 0.554 | 0.92 | 0.72 | 1.18 | 0.569 |
| ≥ X̄+1SD | 24268 | 56 | 1.41 | 1.00 | 1.99 | 0.141 | 1.45 | 1.03 | 2.04 | 0.141 | 1.51 | 1.07 | 2.13 | 0.141 |
| Dementia in other diseases | | | | | | | | | | | | | | |
| ≤ X̄-1SD | 31566 | 79 | References | | | | | | | | | | | |
| Middle | 120644 | 209 | 0.73 | 0.56 | 0.95 | 0.141 | 0.73 | 0.56 | 0.95 | 0.141 | 0.74 | 0.57 | 0.96 | 0.141 |
| ≥ X̄+1SD | 24235 | 23 | 0.60 | 0.38 | 0.96 | 0.141 | 0.60 | 0.38 | 0.96 | 0.141 | 0.63 | 0.39 | 1.00 | 0.141 |
| Dementia (unspecified) | | | | | | | | | | | | | | |
| ≤ X̄-1SD | 31736 | 249 | References | | | | | | | | | | | |
| Middle | 121326 | 891 | 0.92 | 0.80 | 1.06 | 0.403 | 0.94 | 0.81 | 1.08 | 0.461 | 0.95 | 0.83 | 1.10 | 0.569 |
| ≥ X̄+1SD | 24353 | 141 | 1.10 | 0.89 | 1.36 | 0.461 | 1.13 | 0.92 | 1.40 | 0.403 | 1.19 | 0.97 | 1.47 | 0.225 |
| All-cause dementia | | | | | | | | | | | | | | |
| ≤ X̄-1SD | 31935 | 448 | References | | | | | | | | | | | |
| Middle | 122014 | 1579 | 0.90 | 0.81 | 1.00 | 0.141 | 0.91 | 0.82 | 1.02 | 0.225 | 0.93 | 0.84 | 1.03 | 0.365 |
| ≥ X̄+1SD | 24468 | 256 | 1.09 | 0.93 | 1.28 | 0.403 | 1.11 | 0.95 | 1.30 | 0.365 | 1.17 | 1.00 | 1.37 | 0.141 |

*Note:* HR = hazard ratio; CI = confidence interval; SD = standard deviation. Time since the baseline assessment (in days) was used as the underlying time axis. Model 1–adjusted for age and sex; Model 2–adjusted for age, sex, highest educational/professional qualification, cohabitation with spouse/partner, annual gross household income, Townsend deprivation index and fasting time; Model 2B–additionally adjusted for apolipoprotein E (*APOE*) genotype. *P*-values shown are corrected for multiple testing using the Benjamini–Hochberg procedure (across all outcomes, exposure levels and models).

#### 15. Descriptive statistics dementia age of onset

**Table S12.** Dementia age of onset

| | *N* | Mean | SD |
|---|---|---|---|
| Alzheimer's disease | 1881 | 75.57 | 5.06 |
| Vascular dementia | 933 | 75.37 | 5.05 |
| Dementia in other diseases | 512 | 74.24 | 5.20 |
| Dementia (unspecified) | 2240 | 75.33 | 5.11 |
| All-cause dementia | 3976 | 75.10 | 5.27 |

*Note:* SD = standard deviation.

#### 16. Polygenic scores and MileAge

**Table S13.** Polygenic scores and MileAge delta

| Polygenic score | *β* | 95% CI | | *p* |
|---|---|---|---|---|
| Alzheimer's disease | -0.039 | -0.054 | -0.023 | <0.001 |
| Vascular dementia | 0.006 | -0.010 | 0.022 | 0.677 |
| Dementia in other diseases | -0.016 | -0.032 | 0.000 | 0.125 |
| Dementia (unspecified) | -0.011 | -0.028 | 0.006 | 0.425 |
| All-cause dementia | -0.008 | -0.026 | 0.010 | 0.677 |

*Note:* CI = confidence interval. Models were adjusted for the first six population principal components, genotype batch number, assessment centre, age, sex, highest educational/professional qualification, cohabitation with spouse/partner, annual gross household income, Townsend deprivation index and fasting time. *P*-values shown are corrected for multiple testing using the Benjamini–Hochberg procedure. $N = 219{,}481$.

#### 17. Ancestry-specific polygenic scores and MileAge delta

**Table S14.** Ancestry-specific polygenic scores and MileAge delta

| Polygenic score | Population | *β* | 95% CI | | *p* |
|---|---|---|---|---|---|
| Alzheimer's disease | AFR | -0.033 | -0.178 | 0.112 | 0.800 |
| | AMR | -0.359 | -0.952 | 0.233 | 0.835 |
| | CSA | -0.174 | -0.300 | -0.047 | 0.019 |
| | EAS | 0.363 | 0.096 | 0.630 | 0.052 |
| | EUR | -0.038 | -0.053 | -0.022 | <0.001 |
| | MID | 0.700 | -0.487 | 1.886 | 0.855 |
| Vascular dementia | AFR | 0.065 | -0.079 | 0.209 | 0.800 |
| | AMR | -0.252 | -0.919 | 0.414 | 0.835 |
| | CSA | -0.122 | -0.250 | 0.006 | 0.124 |
| | EAS | 0.249 | -0.034 | 0.531 | 0.155 |
| | EUR | 0.006 | -0.010 | 0.023 | 0.695 |
| | MID | -1.203 | -2.279 | -0.126 | 0.322 |
| Dementia in other diseases | AFR | -0.100 | -0.243 | 0.044 | 0.638 |
| | AMR | 0.125 | -0.477 | 0.727 | 0.847 |
| | CSA | -0.030 | -0.157 | 0.097 | 0.642 |
| | EAS | -0.047 | -0.348 | 0.254 | 0.760 |
| | EUR | -0.015 | -0.031 | 0.002 | 0.222 |
| | MID | -0.493 | -1.689 | 0.703 | 0.855 |
| Dementia (unspecified) | AFR | -0.100 | -0.232 | 0.033 | 0.638 |
| | AMR | -0.321 | -0.987 | 0.345 | 0.835 |
| | CSA | -0.230 | -0.359 | -0.102 | 0.005 |
| | EAS | 0.242 | -0.060 | 0.544 | 0.183 |
| | EUR | -0.008 | -0.025 | 0.010 | 0.695 |
| | MID | 0.135 | -0.821 | 1.091 | 0.855 |
| All-cause dementia | AFR | -0.032 | -0.169 | 0.106 | 0.800 |
| | AMR | -0.273 | -0.946 | 0.401 | 0.835 |
| | CSA | -0.219 | -0.350 | -0.087 | 0.005 |
| | EAS | 0.373 | 0.092 | 0.655 | 0.052 |
| | EUR | -0.006 | -0.025 | 0.012 | 0.713 |
| | MID | -0.177 | -1.339 | 0.985 | 0.855 |

*Note:* CI = confidence interval; AFR = African; AMR = Admixed American; EAS = East Asian; EUR = European; CSA = Central and South Asian; MID = Middle Eastern. Models were adjusted for the first six population principal components, genotype batch number, assessment centre, age, sex, highest educational/professional qualification, cohabitation with spouse/partner, annual gross household income, Townsend deprivation index and fasting time. *P*-values shown are corrected for multiple testing using the Benjamini–Hochberg procedure (across all polygenic scores and populations). *N* = 3426 (AFR); *N* = 278 (AMR); *N* = 3916 (CSA); *N* = 1012 (EAS); *N* = 210,755 (EUR); *N* = 94 (MID).

#### 18. *APOE* genotype and MileAge

**Table S15.** *APOE* genotype and MileAge delta

| | *N* | *β* | 95% CI | | *p* |
|---|---|---|---|---|---|
| *APOE* genotype | | | | | |
| ε2ε2 | 1343 | 0.489 | 0.287 | 0.690 | <0.001 |
| ε2ε3 | 27185 | 0.373 | 0.324 | 0.422 | <0.001 |
| ε3ε3 | 5655 | 0.390 | 0.290 | 0.490 | <0.001 |
| ε2ε4 | 131236 | Reference | | | |
| ε4ε3 | 52719 | 0.024 | -0.014 | 0.062 | 0.260 |
| ε4ε4 | 5358 | 0.031 | -0.072 | 0.134 | 0.552 |
| *APOE* risk categories | | | | | |
| Low | 28528 | 0.379 | 0.330 | 0.427 | <0.001 |
| Average | 131236 | Reference | | | |
| Moderate | 58374 | 0.060 | 0.023 | 0.096 | 0.002 |
| High | 5358 | 0.031 | -0.072 | 0.133 | 0.556 |

*Note: APOE* = apolipoprotein E; CI = confidence interval. Models were adjusted for the first six population principal components, genotype batch number, assessment centre, age, sex, highest educational/professional qualification, cohabitation with spouse/partner, annual gross household income, Townsend deprivation index and fasting time. *P*-values shown are corrected for multiple testing using the Benjamini–Hochberg procedure (across all exposure levels).

#### 19. *APOE* genotype and incident dementia

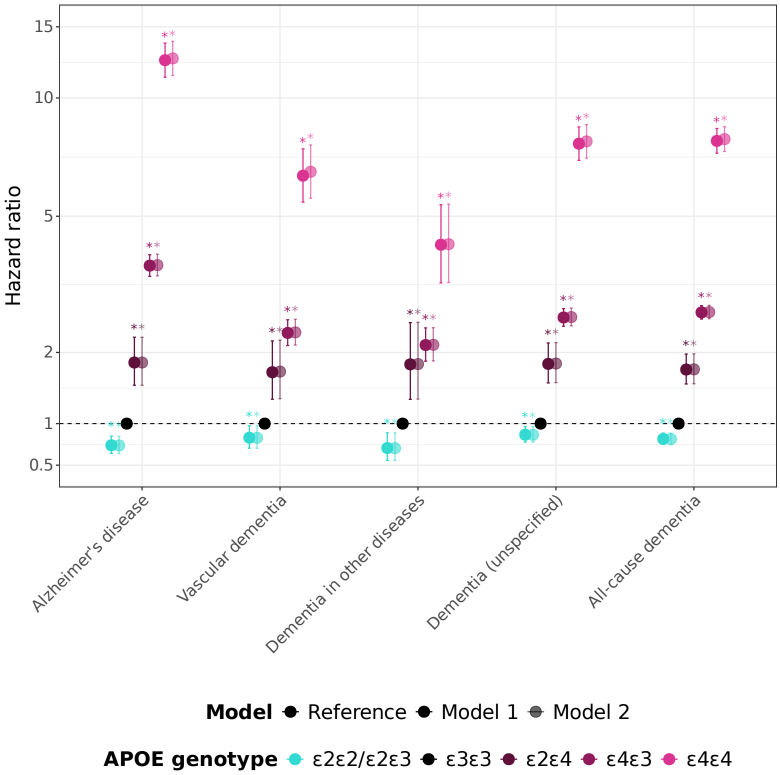

**Figure S9.** *APOE* = apolipoprotein E. Time since the baseline assessment (in days) was used as the underlying time axis. Model 1–adjusted for the first six population principal components, genotype batch number, assessment centre, age and sex; Model 2–adjusted for the first six population principal components, genotype batch number, assessment centre, age, sex, highest educational/professional qualification, cohabitation with spouse/partner, annual gross household income and Townsend deprivation index. Asterisks indicate statistical significance after correcting *p*-values for multiple testing using the Benjamini–Hochberg procedure.

**Table S16.** *APOE* genotype and incident dementia

| | | | Model 1 | | | | Model 2 | | | |
|---|---|---|---|---|---|---|---|---|---|---|
| Level | $N_{\text{total}}$ | $N_{\text{incident}}$ | HR | 95% CI | | *p* | HR | 95% CI | | *p* |
| Alzheimer's disease | | | | | | | | | | |
| ε3ε3 | 283822 | 1384 | Reference | | | | | | | |
| ε2ε2/ε2ε3 | 62167 | 225 | 0.73 | 0.64 | 0.85 | <0.001 | 0.73 | 0.64 | 0.84 | <0.001 |
| ε2ε4 | 12097 | 104 | 1.84 | 1.51 | 2.25 | <0.001 | 1.84 | 1.51 | 2.25 | <0.001 |
| ε4ε3 | 112911 | 1930 | 3.68 | 3.43 | 3.94 | <0.001 | 3.69 | 3.44 | 3.96 | <0.001 |
| ε4ε4 | 11183 | 584 | 12.41 | 11.26 | 13.68 | <0.001 | 12.53 | 11.37 | 13.81 | <0.001 |
| Vascular dementia | | | | | | | | | | |
| ε3ε3 | 283344 | 906 | Reference | | | | | | | |
| ε2ε2/ε2ε3 | 62109 | 167 | 0.83 | 0.70 | 0.97 | 0.023 | 0.82 | 0.70 | 0.97 | 0.022 |
| ε2ε4 | 12057 | 64 | 1.70 | 1.32 | 2.19 | <0.001 | 1.71 | 1.32 | 2.20 | <0.001 |
| ε4ε3 | 111770 | 789 | 2.32 | 2.11 | 2.56 | <0.001 | 2.33 | 2.12 | 2.57 | <0.001 |
| ε4ε4 | 10790 | 191 | 6.37 | 5.44 | 7.45 | <0.001 | 6.52 | 5.57 | 7.63 | <0.001 |
| Dementia in other diseases | | | | | | | | | | |
| ε3ε3 | 282963 | 525 | Reference | | | | | | | |
| ε2ε2/ε2ε3 | 62024 | 82 | 0.70 | 0.56 | 0.89 | 0.003 | 0.70 | 0.55 | 0.88 | 0.003 |
| ε2ε4 | 12033 | 40 | 1.81 | 1.32 | 2.50 | <0.001 | 1.82 | 1.32 | 2.51 | <0.001 |
| ε4ε3 | 111401 | 420 | 2.12 | 1.86 | 2.41 | <0.001 | 2.12 | 1.87 | 2.41 | <0.001 |
| ε4ε4 | 10673 | 74 | 4.20 | 3.29 | 5.36 | <0.001 | 4.21 | 3.30 | 5.38 | <0.001 |
| Dementia (unspecified) | | | | | | | | | | |
| ε3ε3 | 284416 | 1978 | Reference | | | | | | | |
| ε2ε2/ε2ε3 | 62320 | 378 | 0.86 | 0.77 | 0.96 | 0.009 | 0.86 | 0.77 | 0.96 | 0.008 |
| ε2ε4 | 12142 | 149 | 1.82 | 1.54 | 2.15 | <0.001 | 1.83 | 1.55 | 2.16 | <0.001 |
| ε4ε3 | 112922 | 1941 | 2.60 | 2.44 | 2.77 | <0.001 | 2.61 | 2.45 | 2.77 | <0.001 |
| ε4ε4 | 11118 | 519 | 7.68 | 6.97 | 8.46 | <0.001 | 7.78 | 7.06 | 8.58 | <0.001 |
| All-cause dementia | | | | | | | | | | |
| ε3ε3 | 285909 | 3471 | Reference | | | | | | | |
| ε2ε2/ε2ε3 | 62564 | 622 | 0.81 | 0.74 | 0.88 | <0.001 | 0.81 | 0.74 | 0.88 | <0.001 |
| ε2ε4 | 12244 | 251 | 1.74 | 1.53 | 1.97 | <0.001 | 1.74 | 1.53 | 1.98 | <0.001 |
| ε4ε3 | 114569 | 3588 | 2.69 | 2.57 | 2.82 | <0.001 | 2.70 | 2.58 | 2.83 | <0.001 |
| ε4ε4 | 11562 | 963 | 7.81 | 7.27 | 8.39 | <0.001 | 7.89 | 7.34 | 8.47 | <0.001 |

*Note: APOE* = apolipoprotein E; HR = hazard ratio; CI = confidence interval. Time since the baseline assessment (in days) was used as the underlying time axis. Model 1–adjusted for the first six population principal components, genotype batch number, assessment centre, age and sex; Model 2–adjusted for the first six population principal components, genotype batch number, assessment centre, age, sex, highest educational/professional qualification, cohabitation with spouse/partner, annual gross household income and Townsend deprivation index. *P*-values shown are corrected for multiple testing using the Benjamini–Hochberg procedure (across all outcomes, exposure levels and models).

#### 20. MileAge delta and incident dementia by *APOE* genotype

**Table S17.** MileAge delta and incident dementia by *APOE* genotype

| | | | | Model 1 | | | | Model 2 | | | |
|---|---|---|---|---|---|---|---|---|---|---|---|
| *APOE* | Level | $N_{\text{total}}$ | $N_{\text{incident}}$ | HR | 95% CI | | *p* | HR | 95% CI | | *p* |
| Alzheimer's disease | | | | | | | | | | | |
| Low | ≤-1SD | 4073 | 21 | 1.24 | 0.80 | 1.92 | 0.409 | 1.24 | 0.80 | 1.92 | 0.409 |
| | Middle | 19122 | 56 | 0.59 | 0.44 | 0.77 | <0.001 | 0.58 | 0.44 | 0.77 | <0.001 |
| | ≥+1SD | 5162 | 27 | 1.36 | 0.92 | 2.01 | 0.159 | 1.34 | 0.91 | 1.98 | 0.182 |
| Average | ≤-1SD | 21586 | 100 | 1.05 | 0.85 | 1.31 | 0.711 | 1.04 | 0.84 | 1.30 | 0.767 |
| | Middle | 88657 | 433 | Reference | | | | | | | |
| | ≥+1SD | 20061 | 86 | 1.14 | 0.90 | 1.43 | 0.350 | 1.12 | 0.89 | 1.41 | 0.409 |
| Moderate | ≤-1SD | 9159 | 143 | 3.61 | 2.98 | 4.36 | <0.001 | 3.57 | 2.95 | 4.31 | <0.001 |
| | Middle | 39415 | 625 | 3.43 | 3.04 | 3.88 | <0.001 | 3.44 | 3.04 | 3.89 | <0.001 |
| | ≥+1SD | 8973 | 120 | 3.88 | 3.17 | 4.75 | <0.001 | 3.78 | 3.09 | 4.63 | <0.001 |
| High | ≤-1SD | 838 | 37 | 11.36 | 8.12 | 15.89 | <0.001 | 11.54 | 8.24 | 16.16 | <0.001 |
| | Middle | 3551 | 192 | 12.74 | 10.75 | 15.10 | <0.001 | 12.74 | 10.75 | 15.11 | <0.001 |
| | ≥+1SD | 804 | 41 | 17.60 | 12.77 | 24.26 | <0.001 | 17.66 | 12.81 | 24.34 | <0.001 |
| Vascular dementia | | | | | | | | | | | |
| Low | ≤-1SD | 4062 | 10 | 0.94 | 0.50 | 1.76 | 0.886 | 0.93 | 0.50 | 1.76 | 0.886 |
| | Middle | 19109 | 43 | 0.74 | 0.54 | 1.03 | 0.101 | 0.74 | 0.53 | 1.01 | 0.087 |
| | ≥+1SD | 5148 | 13 | 1.10 | 0.63 | 1.92 | 0.796 | 1.08 | 0.62 | 1.88 | 0.860 |
| Average | ≤-1SD | 21546 | 60 | 0.99 | 0.75 | 1.31 | 0.947 | 0.98 | 0.74 | 1.30 | 0.934 |
| | Middle | 88485 | 261 | Reference | | | | | | | |
| | ≥+1SD | 20052 | 77 | 1.74 | 1.35 | 2.24 | <0.001 | 1.71 | 1.33 | 2.21 | <0.001 |
| Moderate | ≤-1SD | 9071 | 55 | 2.21 | 1.65 | 2.96 | <0.001 | 2.18 | 1.63 | 2.92 | <0.001 |
| | Middle | 39057 | 267 | 2.46 | 2.08 | 2.92 | <0.001 | 2.46 | 2.07 | 2.92 | <0.001 |
| | ≥+1SD | 8911 | 58 | 3.24 | 2.43 | 4.30 | <0.001 | 3.10 | 2.33 | 4.12 | <0.001 |
| High | ≤-1SD | 811 | 10 | 4.98 | 2.65 | 9.36 | <0.001 | 5.06 | 2.69 | 9.52 | <0.001 |
| | Middle | 3424 | 65 | 7.35 | 5.60 | 9.65 | <0.001 | 7.31 | 5.57 | 9.60 | <0.001 |
| | ≥+1SD | 777 | 14 | 11.21 | 6.55 | 19.21 | <0.001 | 11.43 | 6.67 | 19.60 | <0.001 |
| Dementia in other diseases | | | | | | | | | | | |
| Low | ≤-1SD | 4066 | 14 | 2.00 | 1.16 | 3.45 | 0.020 | 1.98 | 1.15 | 3.42 | 0.022 |
| | Middle | 19089 | 23 | 0.63 | 0.41 | 0.98 | 0.057 | 0.63 | 0.41 | 0.97 | 0.054 |
| | ≥+1SD | 5141 | 6 | 0.77 | 0.34 | 1.73 | 0.601 | 0.75 | 0.33 | 1.70 | 0.589 |
| Average | ≤-1SD | 21530 | 44 | 1.12 | 0.80 | 1.56 | 0.593 | 1.12 | 0.80 | 1.56 | 0.593 |
| | Middle | 88389 | 165 | Reference | | | | | | | |
| | ≥+1SD | 19999 | 24 | 0.82 | 0.54 | 1.26 | 0.448 | 0.82 | 0.53 | 1.26 | 0.442 |
| Moderate | ≤-1SD | 9063 | 47 | 2.91 | 2.10 | 4.02 | <0.001 | 2.88 | 2.08 | 3.99 | <0.001 |
| | Middle | 38929 | 139 | 2.01 | 1.61 | 2.52 | <0.001 | 2.00 | 1.60 | 2.51 | <0.001 |
| | ≥+1SD | 8875 | 22 | 1.83 | 1.18 | 2.86 | 0.012 | 1.79 | 1.15 | 2.80 | 0.016 |
| High | ≤-1SD | 805 | 4 | 3.00 | 1.11 | 8.10 | 0.045 | 2.96 | 1.10 | 7.99 | 0.048 |
| | Middle | 3379 | 20 | 3.55 | 2.23 | 5.65 | <0.001 | 3.47 | 2.18 | 5.53 | <0.001 |
| | ≥+1SD | 767 | 4 | 4.61 | 1.71 | 12.45 | 0.004 | 4.68 | 1.74 | 12.64 | 0.004 |
| Dementia (unspecified) | | | | | | | | | | | |
| Low | ≤-1SD | 4077 | 25 | 1.00 | 0.67 | 1.49 | 0.984 | 0.99 | 0.67 | 1.48 | 0.984 |
| | Middle | 19162 | 96 | 0.69 | 0.56 | 0.86 | 0.001 | 0.69 | 0.55 | 0.85 | 0.001 |
| | ≥+1SD | 5175 | 40 | 1.38 | 1.00 | 1.90 | 0.067 | 1.35 | 0.98 | 1.86 | 0.090 |
| Average | ≤-1SD | 21606 | 120 | 0.85 | 0.70 | 1.03 | 0.136 | 0.84 | 0.69 | 1.02 | 0.118 |
| | Middle | 88853 | 629 | Reference | | | | | | | |
| | ≥+1SD | 20117 | 142 | 1.30 | 1.08 | 1.56 | 0.009 | 1.27 | 1.06 | 1.53 | 0.015 |
| Moderate | ≤-1SD | 9167 | 151 | 2.57 | 2.15 | 3.07 | <0.001 | 2.54 | 2.13 | 3.04 | <0.001 |
| | Middle | 39449 | 659 | 2.49 | 2.24 | 2.78 | <0.001 | 2.50 | 2.24 | 2.79 | <0.001 |
| | ≥+1SD | 8987 | 134 | 2.98 | 2.47 | 3.59 | <0.001 | 2.88 | 2.39 | 3.48 | <0.001 |

| | | | | | | | | | | | |
|---|---|---|---|---|---|---|---|---|---|---|---|
| High | ≤-1SD | 836 | 35 | 7.08 | 5.04 | 9.96 | <0.001 | 7.23 | 5.14 | 10.17 | <0.001 |
| | Middle | 3538 | 179 | 8.15 | 6.90 | 9.62 | <0.001 | 8.12 | 6.88 | 9.60 | <0.001 |
| | ≥+1SD | 793 | 30 | 9.06 | 6.28 | 13.07 | <0.001 | 9.13 | 6.33 | 13.18 | <0.001 |
| All-cause dementia | | | | | | | | | | | |
| Low | ≤-1SD | 4099 | 47 | 1.10 | 0.82 | 1.47 | 0.603 | 1.10 | 0.82 | 1.47 | 0.603 |
| | Middle | 19234 | 168 | 0.71 | 0.61 | 0.84 | <0.001 | 0.71 | 0.60 | 0.83 | <0.001 |
| | ≥+1SD | 5195 | 60 | 1.21 | 0.93 | 1.57 | 0.190 | 1.19 | 0.92 | 1.54 | 0.241 |
| Average | ≤-1SD | 21725 | 239 | 0.99 | 0.86 | 1.14 | 0.934 | 0.99 | 0.86 | 1.13 | 0.886 |
| | Middle | 89295 | 1071 | Reference | | | | | | | |
| | ≥+1SD | 20216 | 241 | 1.28 | 1.12 | 1.48 | <0.001 | 1.27 | 1.10 | 1.46 | 0.002 |
| Moderate | ≤-1SD | 9292 | 276 | 2.71 | 2.37 | 3.09 | <0.001 | 2.68 | 2.35 | 3.06 | <0.001 |
| | Middle | 39986 | 1196 | 2.62 | 2.41 | 2.85 | <0.001 | 2.62 | 2.41 | 2.85 | <0.001 |
| | ≥+1SD | 9096 | 243 | 3.11 | 2.70 | 3.57 | <0.001 | 3.02 | 2.63 | 3.48 | <0.001 |
| High | ≤-1SD | 857 | 56 | 6.55 | 5.01 | 8.57 | <0.001 | 6.66 | 5.09 | 8.72 | <0.001 |
| | Middle | 3677 | 318 | 8.19 | 7.23 | 9.29 | <0.001 | 8.18 | 7.22 | 9.28 | <0.001 |
| | ≥+1SD | 824 | 61 | 10.27 | 7.93 | 13.30 | <0.001 | 10.30 | 7.95 | 13.34 | <0.001 |

*Note:* HR = hazard ratio; CI = confidence interval; SD = standard deviation. Apolipoprotein E (*APOE*) genotypes classified into low (ε2ε2 and ε2ε3), average (ε3ε3), moderate (ε2ε4 and ε4ε3) and high (ε4ε4) risk categories. Time since the baseline assessment (in days) was used as the underlying time axis. Model 1–adjusted for the first six population principal components, age and sex; Model 2–adjusted for the first six population principal components, age, sex, highest educational/professional qualification, cohabitation with spouse/partner and annual gross household income. No adjustment was made for genotype batch number, assessment centre, Townsend deprivation index and fasting time due to sparse data. *P*-values shown are corrected for multiple testing using the Benjamini–Hochberg procedure (across all outcomes, exposure levels and models).

#### 21. MileAge delta and incident dementia by dementia PRS

**Table S18.** MileAge delta and incident dementia by dementia PRS

| | | | | Model 1 | | | | Model 2 | | | |
|---|---|---|---|---|---|---|---|---|---|---|---|
| PRS | Level | $N_{total}$ | $N_{incident}$ | HR | 95% CI | | *p* | HR | 95% CI | | *p* |
| Alzheimer's disease | | | | | | | | | | | |
| Low | ≤-1SD | 5621 | 20 | 0.58 | 0.37 | 0.91 | 0.030 | 0.58 | 0.37 | 0.90 | 0.027 |
| | Middle | 24196 | 83 | 0.51 | 0.40 | 0.64 | <0.001 | 0.51 | 0.40 | 0.64 | <0.001 |
| | ≥+1SD | 5946 | 18 | 0.57 | 0.36 | 0.91 | 0.030 | 0.56 | 0.35 | 0.90 | 0.028 |
| Middle | ≤-1SD | 23983 | 158 | 1.10 | 0.92 | 1.31 | 0.367 | 1.08 | 0.91 | 1.29 | 0.435 |
| | Middle | 100722 | 673 | Reference | | | | | | | |
| | ≥+1SD | 23221 | 160 | 1.34 | 1.13 | 1.60 | 0.002 | 1.32 | 1.11 | 1.57 | 0.003 |
| High | ≤-1SD | 5364 | 121 | 3.94 | 3.25 | 4.79 | <0.001 | 3.97 | 3.27 | 4.82 | <0.001 |
| | Middle | 23162 | 534 | 3.69 | 3.30 | 4.14 | <0.001 | 3.70 | 3.30 | 4.14 | <0.001 |
| | ≥+1SD | 5102 | 91 | 3.92 | 3.14 | 4.88 | <0.001 | 3.86 | 3.10 | 4.81 | <0.001 |
| Vascular dementia | | | | | | | | | | | |
| Low | ≤-1SD | 5706 | 19 | 0.86 | 0.54 | 1.36 | 0.572 | 0.85 | 0.54 | 1.35 | 0.571 |
| | Middle | 24067 | 59 | 0.59 | 0.45 | 0.78 | <0.001 | 0.59 | 0.45 | 0.78 | <0.001 |
| | ≥+1SD | 5502 | 21 | 1.27 | 0.82 | 1.96 | 0.367 | 1.25 | 0.81 | 1.95 | 0.384 |
| Middle | ≤-1SD | 24382 | 85 | 0.88 | 0.70 | 1.11 | 0.367 | 0.87 | 0.69 | 1.10 | 0.343 |
| | Middle | 102798 | 423 | Reference | | | | | | | |
| | ≥+1SD | 23916 | 112 | 1.51 | 1.22 | 1.86 | <0.001 | 1.48 | 1.20 | 1.82 | <0.001 |
| High | ≤-1SD | 4715 | 30 | 1.68 | 1.16 | 2.43 | 0.011 | 1.69 | 1.17 | 2.45 | 0.010 |
| | Middle | 20554 | 147 | 1.81 | 1.50 | 2.18 | <0.001 | 1.79 | 1.49 | 2.16 | <0.001 |
| | ≥+1SD | 4742 | 27 | 1.86 | 1.26 | 2.75 | 0.004 | 1.85 | 1.25 | 2.74 | 0.004 |
| Dementia in other diseases | | | | | | | | | | | |
| Low | ≤-1SD | 6069 | 19 | 1.36 | 0.85 | 2.18 | 0.276 | 1.37 | 0.86 | 2.19 | 0.276 |
| | Middle | 25610 | 44 | 0.70 | 0.51 | 0.97 | 0.052 | 0.71 | 0.51 | 0.98 | 0.055 |
| | ≥+1SD | 5876 | 7 | 0.62 | 0.29 | 1.32 | 0.308 | 0.63 | 0.30 | 1.33 | 0.309 |
| Middle | ≤-1SD | 23945 | 74 | 1.30 | 1.00 | 1.69 | 0.076 | 1.30 | 1.00 | 1.68 | 0.078 |
| | Middle | 101792 | 247 | Reference | | | | | | | |
| | ≥+1SD | 23619 | 40 | 0.90 | 0.64 | 1.25 | 0.584 | 0.89 | 0.64 | 1.25 | 0.571 |
| High | ≤-1SD | 4764 | 16 | 1.41 | 0.85 | 2.35 | 0.272 | 1.40 | 0.84 | 2.33 | 0.276 |
| | Middle | 19732 | 53 | 1.14 | 0.85 | 1.53 | 0.460 | 1.15 | 0.85 | 1.54 | 0.443 |
| | ≥+1SD | 4560 | 8 | 0.95 | 0.47 | 1.93 | 0.917 | 0.95 | 0.47 | 1.92 | 0.913 |
| Dementia (unspecified) | | | | | | | | | | | |
| Low | ≤-1SD | 5250 | 24 | 0.51 | 0.34 | 0.77 | 0.003 | 0.51 | 0.34 | 0.76 | 0.003 |
| | Middle | 22132 | 138 | 0.64 | 0.54 | 0.76 | <0.001 | 0.64 | 0.54 | 0.77 | <0.001 |
| | ≥+1SD | 5212 | 37 | 0.99 | 0.72 | 1.38 | 0.969 | 0.97 | 0.70 | 1.35 | 0.913 |
| Middle | ≤-1SD | 25241 | 226 | 1.02 | 0.89 | 1.18 | 0.803 | 1.01 | 0.87 | 1.17 | 0.917 |
| | Middle | 106991 | 1012 | Reference | | | | | | | |
| | ≥+1SD | 24854 | 228 | 1.25 | 1.09 | 1.45 | 0.004 | 1.23 | 1.07 | 1.42 | 0.009 |
| High | ≤-1SD | 4506 | 78 | 2.05 | 1.63 | 2.59 | <0.001 | 2.04 | 1.62 | 2.57 | <0.001 |
| | Middle | 19206 | 389 | 2.26 | 2.01 | 2.55 | <0.001 | 2.25 | 2.00 | 2.53 | <0.001 |
| | ≥+1SD | 4273 | 74 | 2.53 | 1.99 | 3.20 | <0.001 | 2.50 | 1.97 | 3.17 | <0.001 |
| All-cause dementia | | | | | | | | | | | |
| Low | ≤-1SD | 4862 | 47 | 0.64 | 0.48 | 0.86 | 0.005 | 0.64 | 0.48 | 0.86 | 0.005 |
| | Middle | 20246 | 194 | 0.57 | 0.49 | 0.66 | <0.001 | 0.57 | 0.49 | 0.66 | <0.001 |
| | ≥+1SD | 4666 | 51 | 0.86 | 0.65 | 1.14 | 0.367 | 0.85 | 0.64 | 1.12 | 0.330 |
| Middle | ≤-1SD | 26449 | 412 | 1.02 | 0.92 | 1.14 | 0.768 | 1.01 | 0.91 | 1.12 | 0.913 |
| | Middle | 112464 | 1852 | Reference | | | | | | | |
| | ≥+1SD | 26176 | 423 | 1.26 | 1.13 | 1.40 | <0.001 | 1.24 | 1.12 | 1.38 | <0.001 |
| High | ≤-1SD | 3971 | 154 | 2.60 | 2.20 | 3.07 | <0.001 | 2.61 | 2.21 | 3.08 | <0.001 |
| | Middle | 16800 | 674 | 2.59 | 2.37 | 2.83 | <0.001 | 2.58 | 2.37 | 2.82 | <0.001 |

| | | | | | | | | | |
|---|---|---|---|---|---|---|---|---|---|
| | ≥+1SD | 3752 | 120 | 2.79 | 2.32 | 3.36 | <0.001 | 2.78 | 2.31 | 3.35 | <0.001 |

*Note:* PRS = polygenic scores; HR = hazard ratio; CI = confidence interval; SD = standard deviation. Polygenic scores classified into low (≤ -1SD), middle and high (≥ +1SD) risk categories. Time since the baseline assessment (in days) was used as the underlying time axis. Model 1–adjusted for the first six population principal components, genotype batch number, assessment centre, age and sex; Model 2–adjusted for the first six population principal components, genotype batch number, assessment centre, age, sex, highest educational/professional qualification, cohabitation with spouse/partner, annual gross household income, Townsend deprivation index and fasting time. *P*-values shown are corrected for multiple testing using the Benjamini–Hochberg procedure (across all outcomes, exposure levels and models).

#### 22. Cross-product interactions MileAge delta and *APOE* genotype

**Table S19.** Cross-product interactions MileAge delta and *APOE* genotype

| | Model 1 | | | | Model 2 | | | |
|---|---|---|---|---|---|---|---|---|
| *APOE* | HR | 95% CI | | *p* | HR | 95% CI | | *p* |
| Alzheimer's disease | | | | | | | | |
| Average | 1.00 | 0.94 | 1.06 | 0.992 | 1.00 | 0.95 | 1.07 | 0.992 |
| Moderate | 1.00 | 0.94 | 1.06 | 0.992 | 1.00 | 0.95 | 1.06 | 0.992 |
| High | 1.02 | 0.96 | 1.09 | 0.992 | 1.02 | 0.96 | 1.09 | 0.992 |
| Vascular dementia | | | | | | | | |
| Average | 1.03 | 0.95 | 1.11 | 0.992 | 1.03 | 0.95 | 1.11 | 0.992 |
| Moderate | 1.01 | 0.94 | 1.09 | 0.992 | 1.01 | 0.94 | 1.09 | 0.992 |
| High | 1.00 | 0.91 | 1.10 | 0.992 | 1.00 | 0.92 | 1.10 | 0.992 |
| Dementia in other diseases | | | | | | | | |
| Average | 1.06 | 0.97 | 1.16 | 0.992 | 1.06 | 0.97 | 1.16 | 0.992 |
| Moderate | 1.05 | 0.96 | 1.15 | 0.992 | 1.05 | 0.96 | 1.15 | 0.992 |
| High | 1.16 | 1.02 | 1.33 | 0.415 | 1.17 | 1.02 | 1.33 | 0.415 |
| Dementia (unspecified) | | | | | | | | |
| Average | 0.99 | 0.94 | 1.04 | 0.992 | 0.99 | 0.95 | 1.04 | 0.992 |
| Moderate | 0.97 | 0.93 | 1.02 | 0.992 | 0.98 | 0.93 | 1.02 | 0.992 |
| High | 0.99 | 0.93 | 1.04 | 0.992 | 0.99 | 0.93 | 1.05 | 0.992 |
| All-cause dementia | | | | | | | | |
| Average | 1.00 | 0.97 | 1.04 | 0.992 | 1.01 | 0.97 | 1.04 | 0.992 |
| Moderate | 1.00 | 0.96 | 1.03 | 0.992 | 1.00 | 0.96 | 1.04 | 0.992 |
| High | 1.02 | 0.97 | 1.06 | 0.992 | 1.02 | 0.97 | 1.06 | 0.992 |

*Note: APOE* = apolipoprotein E; HR = hazard ratio; CI = confidence interval. Time since the baseline assessment (in days) was used as the underlying time axis. Model 1–adjusted for the first six population principal components, genotype batch number, assessment centre, age and sex; Model 2–adjusted for the first six population principal components, genotype batch number, assessment centre, age, sex, highest educational/professional qualification, cohabitation with spouse/partner, annual gross household income, Townsend deprivation index and fasting time. *P*-values shown are corrected for multiple testing using the Benjamini–Hochberg procedure (across all outcomes, exposure levels and models).

#### 23. Cross-product interactions MileAge delta and dementia PRS

**Table S20.** Cross-product interactions MileAge delta and dementia PRS

| | Model 1 | | | | Model 2 | | | |
|---|---|---|---|---|---|---|---|---|
| Polygenic score | HR | 95% CI | | *p* | HR | 95% CI | | *p* |
| Alzheimer's disease | 1.00 | 0.99 | 1.01 | 0.911 | 1.00 | 0.99 | 1.01 | 0.911 |
| Vascular dementia | 1.00 | 0.99 | 1.02 | 0.911 | 1.00 | 0.99 | 1.02 | 0.911 |
| Dementia in other diseases | 1.01 | 0.98 | 1.03 | 0.911 | 1.01 | 0.98 | 1.03 | 0.911 |
| Dementia (unspecified) | 1.00 | 0.99 | 1.01 | 0.911 | 1.00 | 0.99 | 1.01 | 0.911 |
| All-cause dementia | 1.00 | 0.99 | 1.01 | 0.911 | 1.00 | 0.99 | 1.01 | 0.911 |

*Note:* PRS = polygenic score; HR = hazard ratio; CI = confidence interval. Time since the baseline assessment (in days) was used as the underlying time axis. Model 1–adjusted for the first six population principal components, genotype batch number, assessment centre, age and sex; Model 2–adjusted for the first six population principal components, genotype batch number, assessment centre, age, sex, highest educational/professional qualification, cohabitation with spouse/partner, annual gross household income, Townsend deprivation index and fasting time. *P*-values shown are corrected for multiple testing using the Benjamini–Hochberg procedure (across all polygenic scores and models).
